# RISK OF HOSPITALISATION WITH COVID-19 AMONG TEACHERS COMPARED TO HEALTHCARE WORKERS AND OTHER WORKING-AGE ADULTS. A NATIONWIDE CASE-CONTROL STUDY

**DOI:** 10.1101/2021.02.05.21251189

**Authors:** Fenton Lynda, Gribben Ciara, Caldwell David, Colville Sam, Bishop Jen, Reid Martin, White Jane, Marion Campbell, S Hutchinson, C Robertson, M Colhoun Helen, Wood Rachael, M McKeigue Paul, A McAllister David

## Abstract

**Objective:** To determine the risk of hospitalisation with COVID-19 and severe COVID-19 among teachers and their household members, overall and compared to healthcare workers and the general working-age population.

**Design:** Population-based nested case-control study.

**Settings:** Scotland, March 2020 to January 2021. Before and after schools re-opened in early August 2020.

**Participants:** All cases of COVID-19 in Scotland in adults ages 21 to 65 (n = 83,817) and a random sample of controls matched on age, sex and general practice (n = 841,708).

**Exposure:** Individuals identified as actively teaching in a Scottish school by the General Teaching Council for Scotland, and household members of such individuals identified via the Unique Property Reference Number.

**Comparator:** Individuals identified as healthcare workers in Scotland, their household members, and the remaining “general population” of working-age adults.

**Main outcomes:** The primary outcome was hospitalisation with COVID-19 defined in anyone testing positive with COVID-19 in hospital, admitted to hospital within 28 days of a positive test, and/or diagnosed with COVID-19 on discharge from hospital. Severe COVID-19 was defined as individuals admitted to intensive care or dying within 28 days of a positive test or assigned COVID-19 as a cause of death.

**Results:** Most teachers were young (mean age 42), female (80%) and had no underlying conditions (84%). The cumulative incidence (risk) of hospitalisation with COVID-19 was below 1% for all of the working age adults. In the period after school re-opening, compared to the general population, in conditional logistic regression models adjusting for age, sex, general practice, deprivation, underlying conditions and number of adults in the household, the relative risk in teachers (among 18,479 cases and controls) for hospitalisation was rate ratio (RR) 0.97 (95%CI 0.72-1.29) and for severe COVID-19 was RR 0.27 (95%CI 0.09-0.77). Equivalent rate ratios for household members of teachers were 0.97 (95%CI 0.74-1.27) and 0.67 (95%CI 0.36-1.24), and for healthcare workers were 1.82 (95%CI 1.55-2.14) and 1.76 (95%CI 1.22-2.56), respectively.

**Conclusion:** Compared to working-age adults who are otherwise similar, teachers and their household members are not at increased risk of hospitalisation with COVID-19 and are at lower risk of severe COVID-19. These findings are broadly reassuring for adults engaged in face to face teaching.

## Introduction

School closures have formed part of the response to the COVID-19 pandemic in nearly every country in the world. While the duration and extent of closures have varied, as of January 2021 children and young people across many countries and regions have no access to schools.^1^ Limiting children’s access to school reduces educational opportunities, limits social interactions and harms physical and mental health, particularly among children from socio-economically deprived backgrounds.^2^

Worldwide, governments have been required to weigh the risks and benefits of school closures. Among the complex considerations are whether providing education in-person poses an increased risk to teachers. This issue has also been a concern for teachers’ representatives. Studies of this risk have been limited by small numbers of events, selection biases and/or a lack of data on potentially important potential confounders such as the prevalence of underlying conditions, or have narrowly focused on specific settings with uncertain wider applicability.^3–7^

Therefore, we aimed to harness the well-established COVID-19 health records informatics infrastructure in Scotland^8–10^ to examine risk of COVID-19 in teachers in Scotland. Although schools in Scotland closed during the first wave of the COVID-19 pandemic, they remained open in the period from August to December 2020, when there was a second wave of infections, and at this time mask-wearing among children was confined to classrooms in secondary schools where older children (aged 12-17) are taught, and only in geographic areas with high rates.^11^

This combination of circumstances provided us with an opportunity to estimate the risk of COVID-19 among teachers in Scotland. Prior to obtaining exposure data, hospitalisation with COVID-19 was pre-specified as the primary outcome. Hospitalisation was chosen rather than focusing on any case of COVID-19 or on severe COVID-19 as the former was judged to be highly susceptible to ascertainment bias (since it is affected by both individual behaviour concerning testing as well as access to testing) while the latter was judged likely to have too few events in working-age populations. We examine relative risks for the periods from March to August 2020 before schools reopened and from September 2020 to January 2021 after re-opening.

## Methods

In brief, we linked datasets of all teachers and - as an additional comparator - all health care workers nationally to an existing case control dataset that contains data on all COVID-19 cases in Scotland and matched population controls. The advantage of linking to an existing case-control study was that we could leverage the extensive data processing and cleaning (especially of covariate data) which we had already performed in order to produce results more rapidly. The case-control study uses incidence-density sampling which means that the effect estimates calculated using these data are identical to hazard ratios obtained from an equivalent whole-population cohort study analysed using Cox proportional hazards models.^12^

The complete case data from the case-control study was also used alongside denominator data (not linked to covariates) which include all teachers and healthcare workers (and by subtraction from the population mid-year estimates, adults in neither category) to allow us to estimate absolute risks in all three groups. The methods are detailed further below.

### Case-control study

Public Health Scotland maintains an incidence-density case-control study sampled from population-wide healthcare utilisation databases held by the organisation. This study, described in detail elsewhere,^9^ includes all individuals in Scotland who are “cases” of COVID-19 (see case definition below), and for each case ten controls randomly selected from the Scottish population who are of the same age (in single years) and sex, and are registered at the same general practice as the case, but who did not (on or before that date) themselves meet the case definition. The case-control study is regularly updated, with the most recent update being performed on the 4^th^ of January 2021. Controls were ascertained using the Community Health Index database which contains the unique health care identifier, other identifiers, age and sex and general practitioner for the total population of Scotland. For the entire analysis, only working aged individuals (those aged 21 to 65 years old) were included. The case-control dataset is linked to recent hospitalisation and prescribing data to identify underlying diseases and to contemporaneous hospitalisation and intensive care data to characterise the severity of each case. Ten controls per case were used.^13^

### Outcomes

All events from the onset of the pandemic until the 4^th^ of January 2021 were included in the analysis.

As in previous analyses, “any case” of COVID-19 was defined as anyone with a positive PCR test for SARS-CoV-2, or a hospital discharge with a diagnosis of COVID-19 regardless of testing positive and/or any death where COVID-19 was included as a cause (regardless of whether it was recorded as the underlying cause and regardless of any prior test).

The primary outcome of this analysis was pre-specified as hospitalisation with COVID-19, defined in anyone with a positive test for SARS-CoV-2 obtained while they were in hospital, or if they were admitted to hospital 28 days or less following a positive test, or if they had a diagnosis of COVID-19 noted on a hospital discharge letter.

The number of teachers and healthcare workers with severe COVID-19 was expected to be low because this outcome is rare among working-age adults and the number of teachers and healthcare workers is relatively small. As such, despite being more robust to clinical decision-making than hospitalisation, severe COVID-19 was selected as a secondary outcome. Severe COVID-19 was defined as COVID-19 resulting in death or admission to intensive care.

### Exposure

The General Teaching Council for Scotland (GTCS) hold data on each teacher including their name, sex, date of birth, home postcode, sector (e.g. nursery, primary or secondary), last known employer and registration number. Teachers are prompted to update their registration details annually. Teachers were defined as individuals of working-age registered with the GTCS, and currently working, or believed to be currently working, as teachers at a Scottish school. The GTCS indicated which individuals were teaching in February 2020, November 2020 or both (see supplementary methods for additional details). These data were linked to the case-control study using name, sex, date of birth, and home postcode. Healthcare workers were identified using the General Practitioner Contractor Database (GPCD) and Scottish Workforce Information Standard System (SWISS) databases as previously described.^10^

Outcomes in teachers were compared simultaneously to both healthcare workers (either known to be in patient-facing roles or to have an “undetermined” status as regards whether or not they were in patient-facing roles) and to working-age adults who were neither teachers nor healthcare workers who are known to be at increased risk; hereafter this groups is termed the “general population” comparator.

Schools in Scotland reopened on or shortly after the 12^th^ of August 2020. A priori, a 21 day lag-period was provided to allow for the time between exposure to SARS-CoV-2 and development of disease severe enough to require hospitalisation and results are presented for before (1^st^ Mar to 2^nd^ Sep 2020) and after schools re-opened (3^rd^ September 2020 to 4^th^ Jan 2021), with more fine-grained time-periods shown in the supplementary appendix.

Household members of healthcare workers and teachers were identified via the Unique Property Reference Number (UPRN) which was added to the national GP-registration database register in 2020.

### Covariates

Within the case-control study, covariates were defined as previously described. Briefly, age, sex and Scottish Index of Multiple Deprivation (SIMD, an area based measure of socio-economic deprivation) were obtained from the national GP-registration database, ethnicity was derived via self-report via a range of healthcare utilisation databases (Scottish Morbidity Records (SMR) 01, 02 and 04) and comorbidity was obtained using previous hospitalisation (SMR01) and drug dispensing (PIS) data using the same definitions developed previously.^9,10^ Additionally, the UPRN was used to obtain counts of the numbers of adults (aged over 18 years) in the household of each case and control.

### Statistical analysis

Summary statistics for demographic, socio-economic and clinical characteristics were calculated for teachers, healthcare workers and the remaining working-age population. The control arm of the case-control study is effectively a stratified random sample from the entire Scottish population, where the strata are defined by the age, sex and the general practice within which each individual was registered. As such, if the probability of inclusion is known, the control arm can be used to obtain valid summary statistics for the whole population; this is analogous to the re-weighting used when analysing survey data. To estimate the inclusion probabilities, we obtained age (in single-years), sex, SIMD and health board area-specific counts of the general Scottish population,^14^ and produced similar counts for the control arm of the case control study. The inclusion probability for any individual was then calculated as the latter count divided by the former. We then produced whole-Scotland statistics for all teachers, healthcare workers and the remaining working-age population using the `TableOne`package in R which allows estimation of summary statistics in the presence of stratified sampling (including counts, proportions, means and standard deviations) via inverse probability weighting.^15^

We produced cumulative incidence plots for hospitalisation with COVID-19 for all three groups, stratifying by age and sex. All events for Scotland were obtained via the case arm of the case-control study with the denominators being obtained directly for teachers and healthcare workers, and via subtraction from the mid-year estimates for the remaining working-age population (`population = mid-year estimates - teachers - healthcare workers`).

For all COVID-19 outcomes - hospitalisation, severe and any case, “unadjusted” conditional logistic regression models were fitted. These effect estimates can be interpreted as, and are hereafter referred to as, rate ratios (RRs). “Unadjusted” models were conditional on the matching variables (age, sex, and general practice). “Adjusted” models additionally included terms for potential confounders such as SIMD, ethnicity, the number of underlying conditions, and whether or not the individual shared a household with a healthcare worker. The pre-specified statistical analysis plan is provided in the supplementary materials and analysis code is available at https://github.com/dmcalli2/tchr.

### Institutional review

We obtained approval from the Health and Social Care Public Benefit and Privacy Panel (HSC-PBPP) to link Public Health Scotland records to an extract of the GTCS register in order to permit estimation of the risk of COVID-19 in teachers (2021-0073). The HSC-PBPP provides scrutiny to ensure information governance principles are maintained, and the panel includes public representatives. A COVID-19 Rapid Data Protection Impact Assessment (DPIA) was completed and approved by the PHS Data Protection Officer. The GTCS notified all registrants of the proposed sharing of registration data in advance, and provided a period in which registrants could raise an objection.

## Results

The teacher dataset included 66,710 unique individuals. By the 4^th^ of January 2021 18,479 had (as cases or controls) been selected into in the 871,568-person case control study. Of 125,830 patient-facing and undetermined healthcare workers, 35,461 were selected into the case control study. Table 1 shows characteristics of the teachers and health care workers compared to the general population using reweighted data from controls as described in the methods above. Compared to the general population, teachers and healthcare workers were similar in terms of age and ethnicity, but were more likely to be women, and to have a lower prevalence of comorbidities. Both teachers and healthcare workers were less likely to live in the most deprived quintile for SIMD than were the general population, with a larger difference for teachers. Teachers were predominantly women; even in secondary schools, where there was a higher proportion of men, two thirds of teachers were women.

**Table 1.**
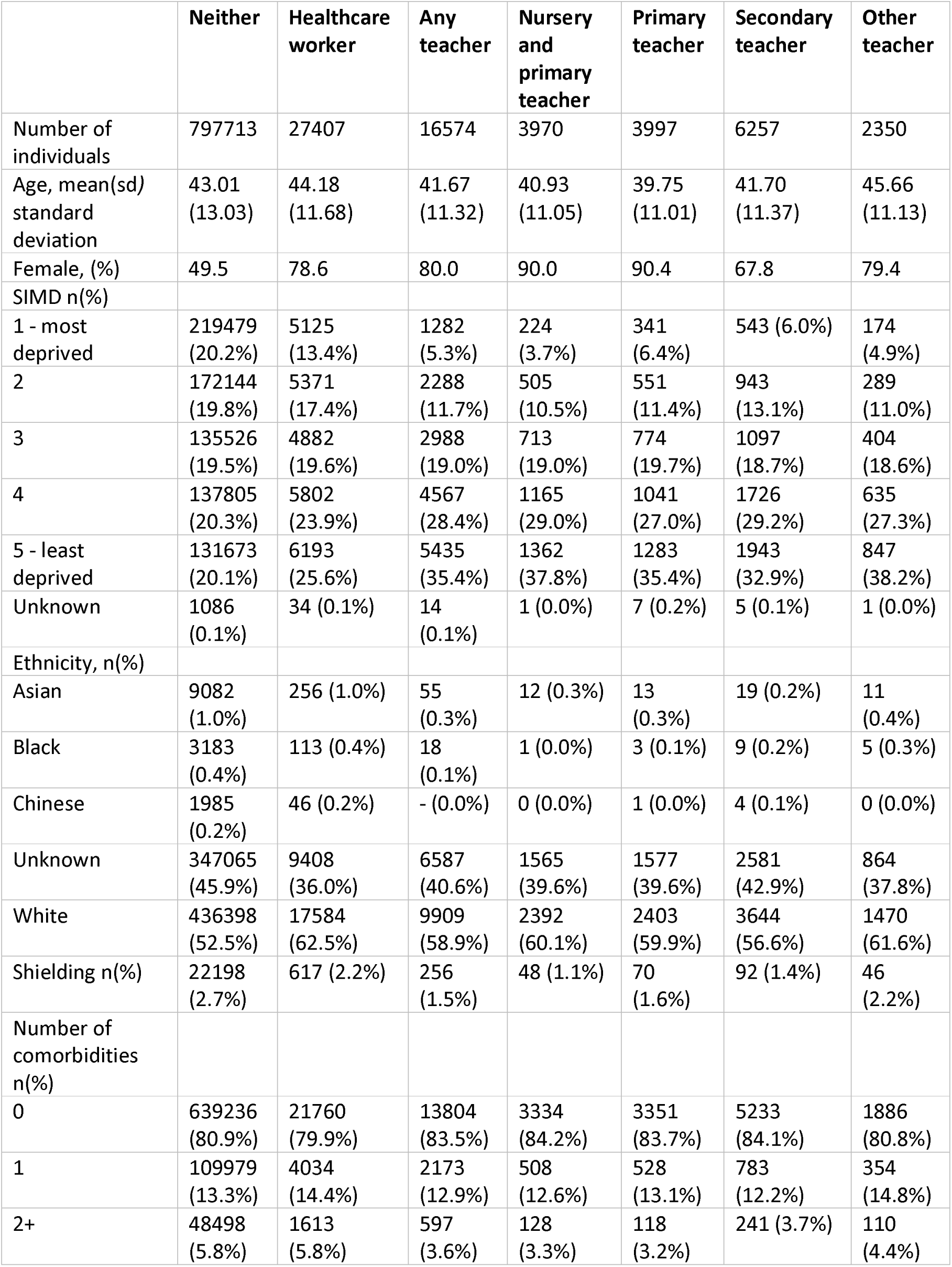
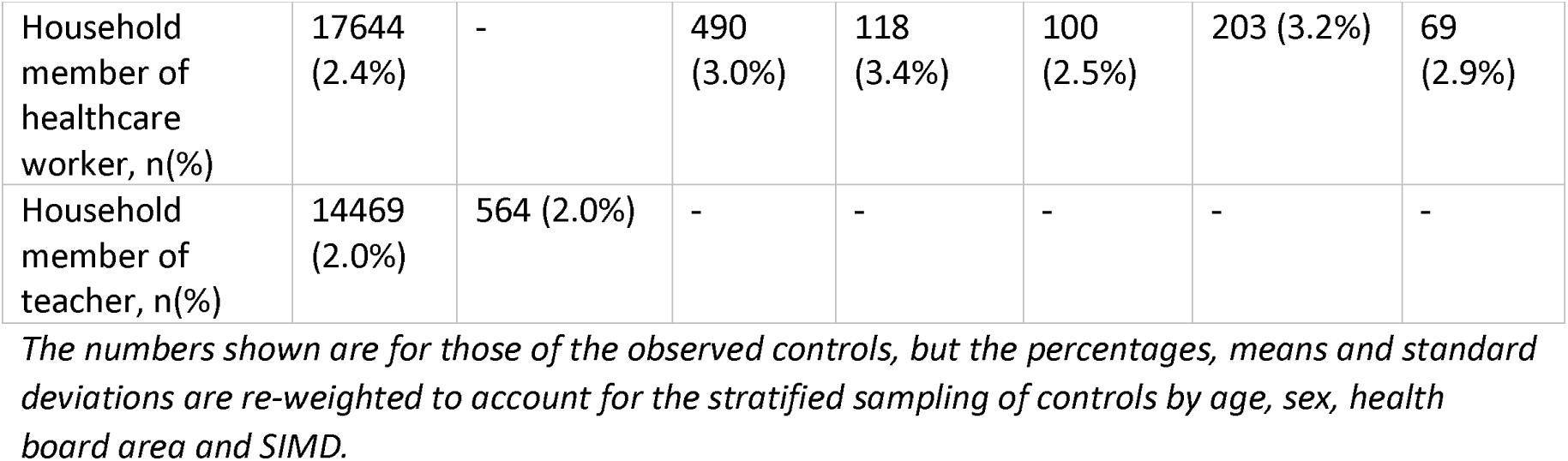
Counts of teachers, healthcare workers and others selected as controls, as well as estimated characteristics of these groups in the Scottish population

### Risks of COVID-19 by occupation

Over the full study period, the cumulative incidence (ie risk) of hospitalisation with COVID-19 has remained below one-percent for teachers, healthcare workers and general population working-age adults (Figure 1). The rise in cumulative incidence over time, and in particular after schools reopened, did not show any clear differences for teachers compared to the other groups.

**Figure 1.**
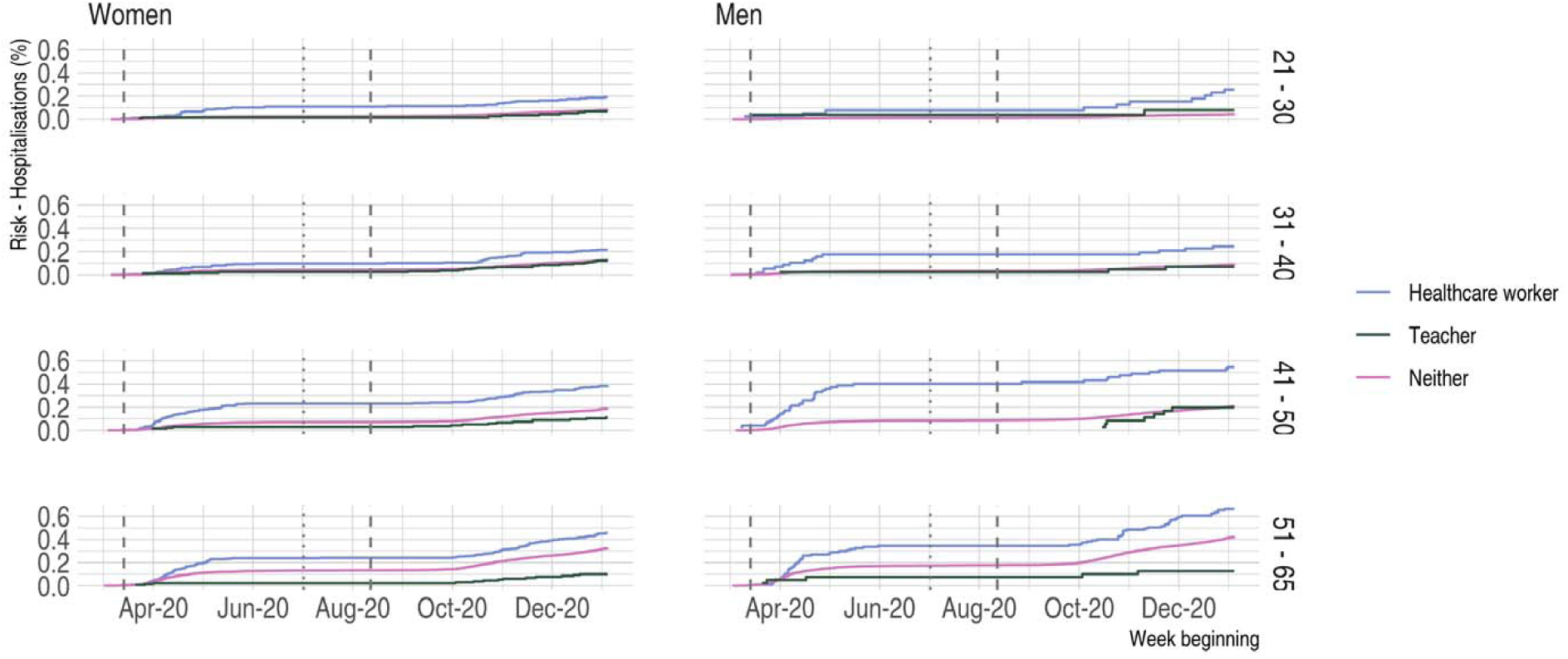
Cumulative incidence (risk) of COVID-19 hospitalisation. The vertical lines indicate the transitions between the following periods - before schools closed, spring/summer restrictions, summer/autumn restrictions, schools re-opened.

### Primary endpoint - hospitalisation

In the period before schools re-opened, teachers and household members of teachers were at lower risk of hospitalisation with COVID-19 than healthcare workers, household members of healthcare workers and the general population; compared to the general population the RRs were 0.51 (95% CI 0.31-0.82) for teachers and 3.15 (95% CI 2.71-3.66) and healthcare workers, and 0.63 (95% CI 0.44-0.91) and 1.87 (95% CI 1.52-2.30) for household members of teachers and healthcare workers respectively (Table 2, Figure 2). Similar results were obtained for the unadjusted and adjusted models (Table 2).

**Table 2.**
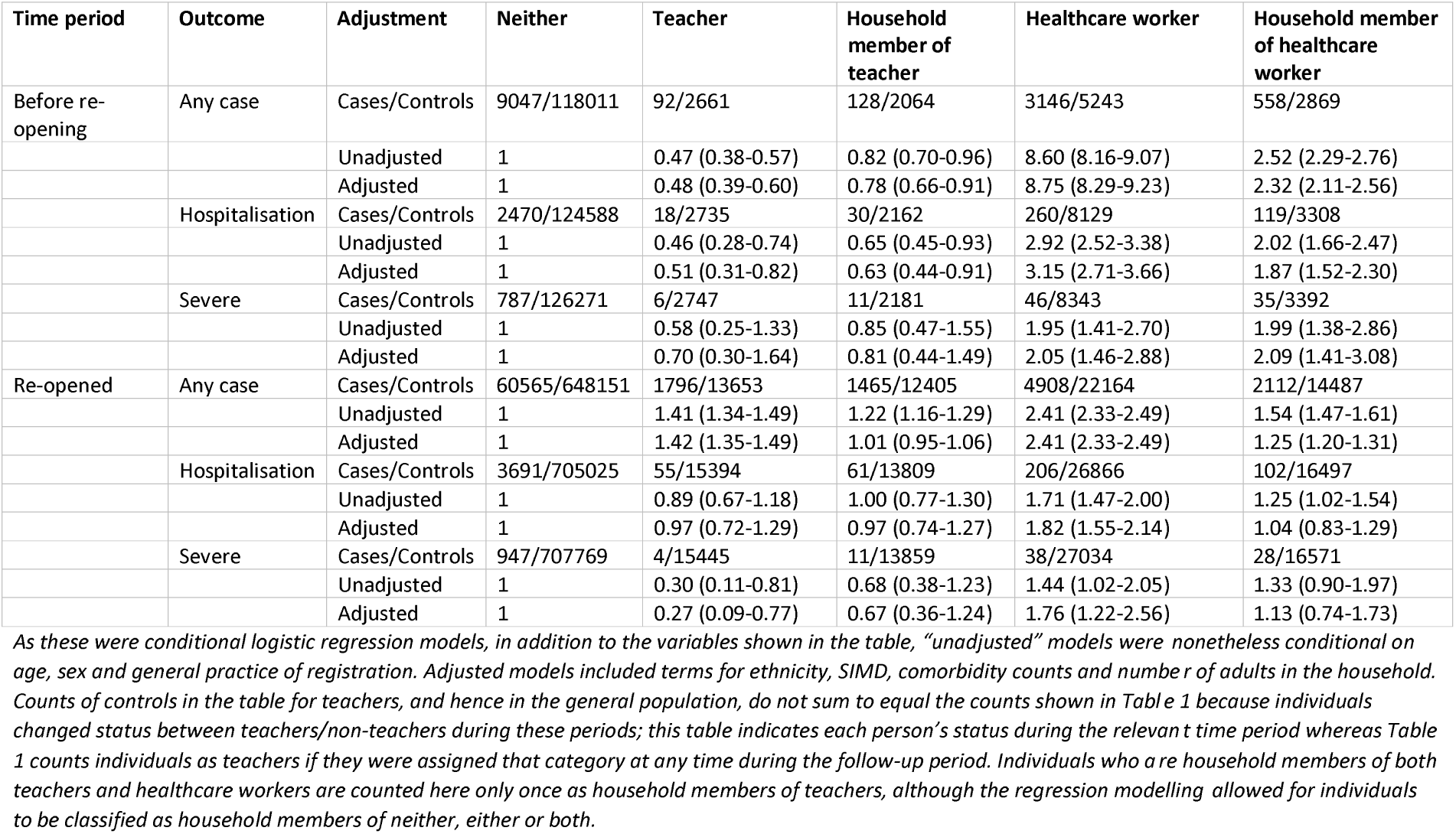
Rate ratios for any case, hospitalisation with COVID-19 and severe COVID-19 for teachers, healthcare workers and members of their households

**Figure 2.**
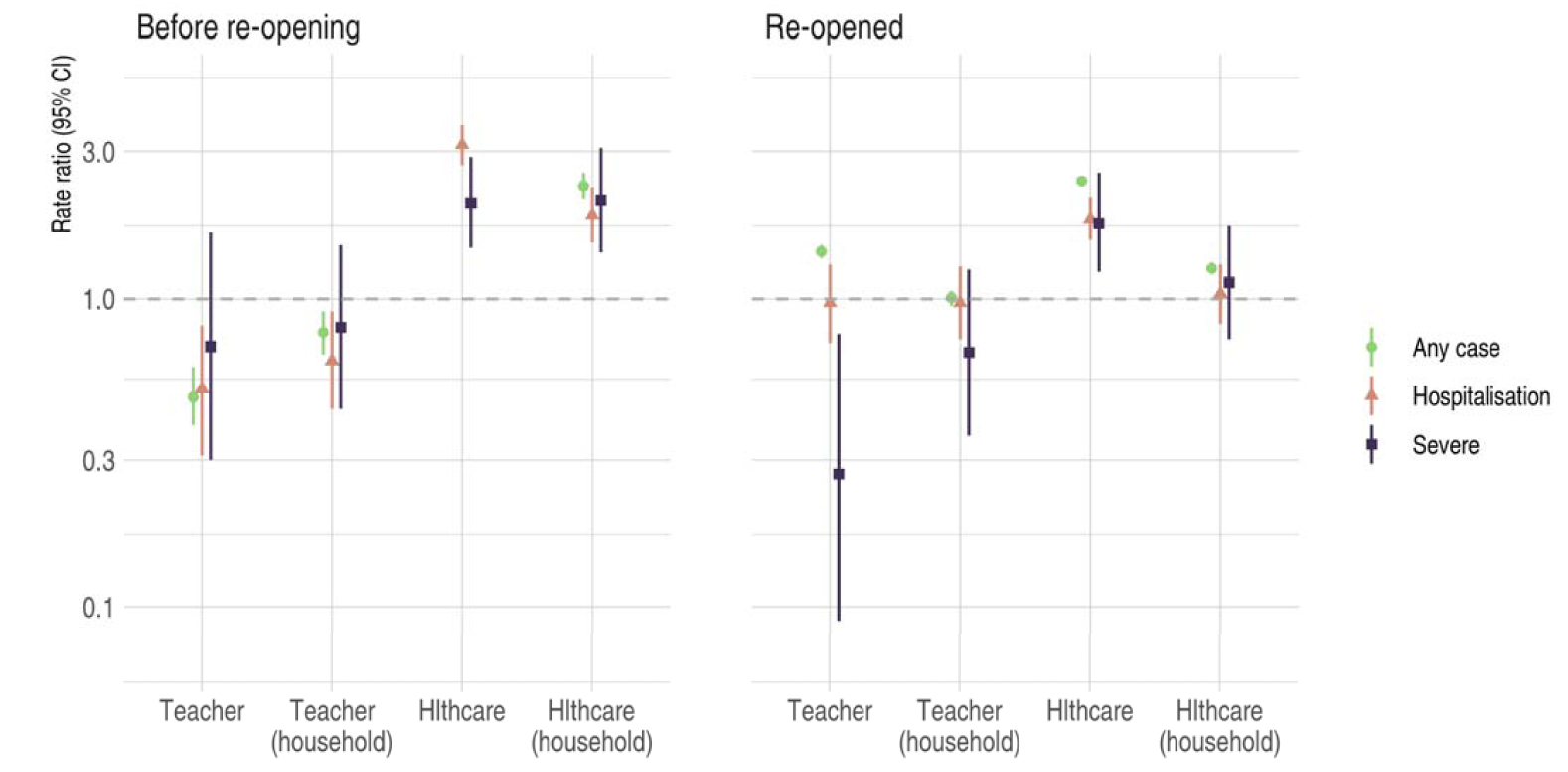
Rate ratios for any case of COVID-19, hospitalisation with COVID-19 and severe COVID-19. Figure represents graphically results shown in Table 2. Points and lines are rate ratios and 95% confidence intervals respectively for adjusted models. The RR for ‘Any case’ among healthcare workers in the period before re-opening is excluded as this was very high making the results for other groups more difficult to distinguish.

In the period after schools re-opened, the rate ratios for hospitalisation in teachers and household members of teachers increased (RR 0.97; 95% CI 0.72-1.29 and 0.97; 95% CI 0.74-1.27 respectively) while RRs for household members of healthcare workers fell (RR 1.04; 95% CI 0.83-1.29) such that they were all closer to one, indicating that all three groups were becoming more similar to the general population. Only healthcare workers had persistently higher rates of hospitalisation than the general population (RR 1.82; 95% CI 1.55-2.14).

### Secondary endpoint – severe COVID-19

For severe COVID-19 the results for healthcare workers and household members of healthcare broadly followed those for hospitalisation with COVID-19 (Table 2, Figure 2). For teachers, however, the RR for severe COVID-19 was markedly lower in the period after schools re-opened; compared to the general population the RR for teachers was 0.27 (95% CI 0.09-0.77).

### Any case of COVID-19

For any case of COVID-19 in the period before schools re-opened teachers had a lower RR compared to the general population (RR 0.48; 95% CI 0.39-0.60), as did household members of teachers (RR 0.78; 95% CI 0.66-0.91). In this period there were higher rates in both healthcare workers (RR 8.75; 95% CI 8.29-9.23) and household members of healthcare workers (RR 2.32; 95% CI 2.11-2.56). In the period after schools re-opened the RRs teachers, healthcare workers and household members of healthcare workers all had higher RRs than the general population, with the highest risk observed among healthcare workers (RRs teachers 1.42; 95% CI 1.35-1.49, healthcare workers 2.41; 95% CI 2.33-2.49 and household members of healthcare workers 1.25; 95% CI 1.20-1.31).There was no difference for household members of teachers (RR 1.01; 95% CI 0.95-1.06). Data on routine testing of teachers and healthcare workers for SARS-CoV-2 is provided in the supplementary appendix.

### Risk of COVID-19 by sector

The rate ratios were broadly similar across different teaching sectors. However, teachers in the “Nursery and Primary” and “Primary” categories (for both categories the overwhelming majority of teachers work with primary-aged children, but the former do so within schools which include a nursery on the same site) had lower rates than secondary teachers and those in the “other” (see supplementary appendix) category. In the period after re-opening, for example, the hospitalisation RRs were lower in both categories of primary teacher (RR 0.61; 95% CI 0.28-1.33 and 0.72; 95% CI 0.37-1.40) than in secondary (RR 1.22; 95% CI 0.81-1.86) or “other” teachers (RR 1.17; 95% CI 0.60-2.27), although in all categories the confidence intervals were wide (Table 3).

**Table 3.** Rate ratios for any case, hospitalisation with COVID-19 and severe COVID-19 for teachers by sector

| Time period | Outcome | Adjustment | Nursery/Primary or Nursery | Primary | Secondary | Other |
| --- | --- | --- | --- | --- | --- | --- |
| Before re-opening | Any case | Counts | 17/672 | 23/641 | 29/970 | 23/378 |
|  |  | Unadjusted | 0.34 (0.21-0.56) | 0.49 (0.32-0.75) | 0.40 (0.28-0.58) | 0.80 (0.52-1.23) |
|  |  | Adjusted | 0.36 (0.22-0.58) | 0.52 (0.34-0.79) | 0.41 (0.28-0.59) | 0.84 (0.55-1.28) |
|  | Hospitalisation | Counts | 2/687 | 4/660 | 8/991 | 4/397 |
|  |  | Unadjusted | 0.27 (0.07-1.12) | 0.42 (0.15-1.14) | 0.52 (0.25-1.06) | 0.57 (0.21-1.59) |
|  |  | Adjusted | 0.30 (0.07-1.23) | 0.48 (0.17-1.31) | 0.56 (0.27-1.15) | 0.65 (0.23-1.84) |
| Re-opened | Any case | Counts | 389/3251 | 501/3304 | 673/5183 | 233/1915 |
|  |  | Unadjusted | 1.30 (1.17-1.44) | 1.64 (1.49-1.80) | 1.38 (1.28-1.50) | 1.30 (1.14-1.49) |
|  |  | Adjusted | 1.30 (1.17-1.45) | 1.66 (1.50-1.83) | 1.39 (1.27-1.51) | 1.31 (1.14-1.51) |
|  | Hospitalisation | Counts | 7/3633 | 10/3795 | 27/5829 | 11/2137 |
|  |  | Unadjusted | 0.57 (0.27-1.22) | 0.73 (0.38-1.38) | 1.06 (0.71-1.58) | 1.08 (0.58-2.01) |
|  |  | Adjusted | 0.61 (0.28-1.33) | 0.72 (0.37-1.40) | 1.22 (0.81-1.86) | 1.17 (0.60-2.27) |

*As these were conditional logistic regression models, in addition to the variables shown in the table, “unadjusted” models were nonetheless conditional on age, sex and general practice of registration. Adjusted models included terms for ethnicity, SIMD, comorbidity counts and number of adults in the household.*

Event numbers and effect estimates from regression models of each outcome by age, sex and more granular time periods are provided in the supplementary appendix and analysis code is available at the project’s GitHub repository.

## Discussion

Among all working-age adults in Scotland, for the period from the first COVID-19 case to the 4^th^ of January 2021, we examined the risks of COVID-19 in teachers and their household members. Neither was at increased risk of hospitalisation or severe COVID-19 at any time, whether compared to healthcare workers or to the general population; both before and after schools re-opened. Teachers were at lower risk of severe COVID-19 (intensive care unit admission or death) than the general population, healthcare workers, and the household members of healthcare workers, and in the period after schools re-opened this difference in severe COVID-19 was statistically significant. These findings were robust to adjustment for potential confounders.

Several previous studies have examined risks of COVID-19 among adults working with children. A US survey of child-care providers (69% response rate) found, on adjusting for age, sex, race, access to personal protective equipment, household income and community rates of infection, that there was no evidence of an association between exposure to child care and risk of COVID-19 (odds ratio (OR) 1.06; 95% CI 0.82-1.38).^16^ A Norwegian study, covering periods where schools were closed and open, provided estimates of the risk of hospitalisation with COVID 19 - adjusting for age, sex and country of birth compared to the general working-age population. The overall estimate was higher in early childhood educators (RR 1.86; 95% CI 0.97-3.57), lower in primary school teachers (RR 0.73; 95% CI 0.44-1.22) and null in secondary education teachers (RR 1.01; 95% CI 0.46-2.21), though all confidence intervals were wide. In a period where schools were open (July 18th to Oct 18th) risks of any COVID-19 (SARS-CoV-2 on PCR testing and/or a coded diagnosis) appeared modestly increased in primary school teachers (RR 1.14; 95% CI 0.99-1.32), secondary education teachers (RR 1.10; 95% CI 0.82-1.47) and childcare workers (RR 1.15; 95% CI 1.02-1.29) but not in “early childhood educators” (RR 0.73; 95% CI 0.54-0.99).^3^

In Sweden, where schools remained mostly open, lower secondary teachers (who taught in person) were compared with upper secondary teachers (who did not). For hospitalisation and death with COVID-19, testing positive for SARS-CoV-2, and diagnosis with COVID-19 - on adjusting for age, sex, income and region - the relative risks were around 2-fold higher in the lower secondary than in upper secondary teachers.^5^ However, other analyses providing comparison with non-teaching occupations did not consistently find increased risk. Compared to IT technicians, teachers in Sweden were not at increased risk of mortality from COVID-19 (adjusting for age, sex, country of birth, living in Stockholm, educational attainment and income), although the confidence intervals were wide (RR 0.91; 95% CI 0.55-1.51).^6^ Compared to other occupations, the age-sex adjusted risk of admission to intensive care for COVID-19 was slightly elevated among preschool teachers (RR 1.10; 95% CI 0.49-2.49) and was lower in school teachers (RR 0.43; 95% CI 0.28-0.68).^7^ Therefore, the differences within secondary teachers may be due to a protective effect of working at home rather than a harmful effect of school-settings.

In the UK, the Office for National Statistics examined deaths from COVID-19 from the 9^th^ of March to the 28^th^ of December, finding that teaching and educational professionals had lower age-standardised mortality than “all residents of England and Wales aged 20 to 64 years” both among men (18.4; 95% CI 14.0-23.6 versus 31.4; 95% CI 30.6-32.3, ranked 19^th^ of 24 occupations from highest to lowest mortality) and women (9.8; 95% CI 7.5-12.5 versus 16.8; 95% CI 16.2-17.5, ranked 15^th^ of 20 from highest to lowest mortality).^17^ In a random sample of the UK population, the ONS found that teachers were around 1.1 times more likely to test positive for SARS-CoV-2 compared to other occupations. However, this comparison was not adjusted for age or sex and the confidence intervals were wide.^4,18^ In a non-random design, Public Health Scotland offered SARS-CoV-2 antibody testing to all individuals working in early learning and school settings in Scotland in October and November 2020. 12,171 teachers opted to participate, and the prevalence of positive antibodies among teaching/teaching support staff was 7.1% (95% CI 6.6 – 7.6),^19^ broadly similar to rates found for individuals aged 14 and older in a nationally representative household survey (7.1%; 95% CI 4.6-10.4 in October 2020).^18^

The international and UK-based findings for hospitalisation and death are therefore broadly consistent with our own observations, demonstrating that teachers are not at increased risk of hospitalisation - and are at lower risk of severe COVID-19 (ICU admission or death) - compared to other members of the working-age population. To these previous reports, which generally adjusted for age, sex and socio-economic status, we add evidence that differences in the prevalence of common underlying conditions does not account for the lower risk of severe COVID-19 outcomes among teachers. We also add the important observation that working-age adults who are household members of teachers are not at increased risk compared to members of the general population.

In our study, we also reported on the risk of “any COVID-19” in teachers compared to the general population, finding a 1.4-fold increase in the period after school opening. In all groups, this measure is largely driven by positive tests in people that have not been hospitalised or become severely ill. We had explicitly not pre-specified this as an outcome because it is very subject to ascertainment bias due to unmeasurable variation in testing policies and practices. These factors not only influence the likelihood of testing, but also the timing and circumstances of testing, such as the presence of symptoms, or regular screening tests; we include this outcome here for completeness only. Survey-based methods are likely to be most appropriate to determine the relative risk for teachers of developing a case of COVID-19 insufficiently severe to require hospitalisation. The ONS School Infection Survey may provide additional information, although with fewer than 5,000 adult participants it will be unable to report on the risks of hospitalisation with COVID-19, which was the focus of our analysis.^20^

Nonetheless, where there is universal health coverage, as in the UK, hospitalisation rates should have considerably less ascertainment bias, while rates of severe COVID-19 should have very little. As such, given that hospitalisation rates were not elevated and that severe disease rates were significantly lower than the background population, if teachers are found to have increased rates of infection one would have to posit some very large effect preferentially preventing infected teachers from becoming seriously ill. Differences in age, sex, calendar time, geography, socio-economic status, underlying conditions or household composition cannot account for any such difference, since these were accounted for in the analysis.

An important finding from our study was the lack of elevation in risks in household members of teachers. This is consistent with a recent review by the European Centre for Disease Control, based largely on contact tracing studies and analyses of outbreaks, which suggested that transmission in schools is relatively uncommon.^21^ In a previous analysis of adults in the household of healthcare workers, we found that (on adjusting for age, sex, SIMD, occupational factors, household factors and underlying conditions) those adults sharing a household with children aged 0-11 years old had a slightly lower risk of testing positive with COVID-19 (HR per child 0.93; 95% CI 0.88-0.98) with similar results (with wider 95% confidence intervals) for the risk of hospitalisation with COVID-19 (HR 0.93; 95% CI 0.79-1.10).^22^ Similarly, the OpenSAFELY study found that in 9 million adults aged 65 or under, sharing a household with children aged 0-11 years was not associated with increased risk of recorded SARS-CoV-2 infection, hospitalisation, ICU admission or death, with similar findings for adults aged over 65.^23^ OpenSAFELY also reported associations per number of child aged 12-17, with broadly similar results. Both the ECDC report, and our findings for household members of children are consistent with our findings for teachers.

A number of studies have examined the impact of closing schools on community transmission of COVID-19, with a recent systematic review of ten studies reporting that the results were equivocal due to difficulties separating the effect of school closures from other policy interventions enacted contemporaneously.^24^ Our study was not designed to examine this question, and in particular cannot be used to examine the indirect impact of school closures on community transmission (eg by signalling to the public that transmission is currently dangerously high thereby increasing the adherence to other non-pharmaceutical interventions).

### Limitations

This was a large and largely complete sample of teachers, and crucially the outcome data was obtained in the same manner for the different occupational groups such that valid comparisons can be made between them, especially for “harder” outcomes such as hospitalisation and admission to intensive care. Nonetheless, there are a number of limitations. First, a small number of GTCS registrants could not be linked to healthcare records. Secondly, the incidence and therefore precision of the estimates was low for hospitalisation and severe COVID-19. Thirdly, the variant of concern (VOC) mutation^25^ did not become dominant in Scotland until mid-December to January, and so we do not yet know how applicable these findings are to this mutation.

Finally, it is important to note that, although the relative risks to teachers was low, as an occupational group, some teachers will nonetheless be at high absolute risk of severe COVID-19 due to combinations of other risk factors such as older age, male sex and underlying conditions. Such individuals should not be considered low risk by virtue of being teachers. Nonetheless since, as we found, most teachers (and to a lesser extent healthcare workers) were young, female, lived in less deprived areas and had no known underlying conditions, the majority of teachers will be at low absolute risk of severe COVID-19 and hospitalisation with COVID-19.

### Conclusion

The majority of teachers were young, female and had few comorbidities and are so at low absolute risk of severe COVID-19 and hospitalisation with COVID-19. Furthermore, compared to working-age adults who are otherwise similar, teachers are not at increased risk of hospitalisation with COVID-19 or severe COVID-19 (and in the case of severe COVID-19 the risk in teachers is actually lower). These findings are reassuring for the majority of adults engaged in face to face teaching.

## Supporting information

Statistical analysis plan

## Data Availability

Data may be accessed via a secure platform following successful application to the Public Benefit and Privacy Panel via application to the electronic Data Research Information Services of Public Health Scotland.

## End matter

### Contribution

LF, MC, SH, CR, HC, RW, PM and DM conceived and designed the study. CG, DC, SC, MR, PM and DM conducted statistical analysis. JW searched the existing literature. DM wrote the first draft and all authors revised the manuscript for intellectual content.

### Patient and public involvement

Patients or the public were not involved in the design, or conduct, or reporting, or dissemination plans of our research.

### Competing interests

All authors have completed the <u>ICMJE uniform disclosure form</u> and declare: no support from any organisation for the submitted work; no financial relationships with any organisations that might have an interest in the submitted work in the previous three years, no other relationships or activities that could appear to have influenced the submitted work.

### Dissemination

Dissemination to study participants and/or patient organisations is not applicable as these results are already known to members of the GTCS.

### Data Sharing

Access to data for third parties would require agreement of the organisations who provided data to Public Health Scotland (NHS Education Scotland and the General Teaching Council Scotland).

## Notes

### Competing Interest Statement

The authors have declared no competing interest.

### Funding Statement

David McAllister is funded via a Wellcome Trust Intermediate Clinical Fellowship and Beit Fellowship (201492/Z/16/Z).

### Author Declarations

Health and Social Care Public Benefit and Privacy Panel

