## Supplementary appendix for "RISK OF HOSPITALISATION WITH COVID-19 AMONG TEACHERS COMPARED TO HEALTHCARE WORKERS AND OTHER WORKING-AGE ADULTS. A NATIONWIDE CASE-CONTROL STUDY"

### Exposure status and the GTCS register

All teachers working in the state sector in Scotland are included on the General Teaching Council for Scotland (GTCS) register, and from 1st October 2017 all newly appointed teachers working at independent schools have also been required to register. However, the deadline for registration for those already in post prior to the 1st of October 2017 is not until 1st of June 2021, so some teachers working in independent schools will not have been included (https://www.gtcs.org.uk/registration/independent-schools-registration.aspx). Teachers were defined by the GTCS as actively teaching if they had completed their “professional update” (the ongoing professional learning requirement that is a requirement for employment as a teacher) and had a last known employer suggestive of a teaching post. A small set of teachers (< 400) were removed from the data before it was provided to PHS at their own request following a consultation held by the GTCS.

Since case/control status is defined at a specific date (generally the date at which a case tested positive for both cases and their matched control), teachers believed **not** to be actively teaching at this time-point were not defined as teachers for this analysis; within the case-control study, 44 individuals were not actively teaching in February but were in November, and 309 teachers were actively teaching in February but not in November. Teachers were further sub-divided based on their self-reported setting. The original settings were collapsed into the following sectors; nursery and primary, primary, secondary and other. Few primary teachers in Scotland are based in nurseries. As such, the majority of teachers allocated to the “nursery and primary” category are teachers of primary-school-aged children (based in schools with on-site nurseries) rather than teachers of pre-school children working within nurseries. The “other” group comprised those working in further education colleges, local government, “miscellaneous”, “nursery/primary/special”, “primary/special” and “special”.

From a total of more than 70,000 registered teachers, the GTCS supplied to PHS 67, 091 records of those believed to be currently working at a school in Scotland. Following de-duplication and the exclusion of records which could not be linked to a CHI-number, the eventual teacher dataset included 66,710 unique individuals.

### Rates of testing and testing positive

While we had results for all teachers who tested positive (via the case arm of the case control study), we did not have negative testing data for all teachers, only for those in the control arm. As such, we estimated the number of teachers tested as follows. For any teacher selected as a control at any time (a stratified random sample of the population), which includes all those who subsequently went on to have a positive test, we excluded all subsequent tests after their first positive result. Next we estimated the proportion of unique individuals tested during each calendar week, by age and sex. Next we applied these proportions to the denominator teacher dataset to estimate the total number of teachers tested each week within each age and sex defined stratum, then summed this to obtain the total number of teachers tested each week. We repeated this exercise for healthcare workers and the general population. Figure S1-A shows the estimated proportion of each group tested using this approach.

Subsequently, we used the case arm of the study to identify the first positive test of each teacher and summed this by age and sex. We then divided this number by the total number in each group to obtain the test rates (Figure S1-B) and by the estimated number tested in each group to obtain the proportion of individuals testing positive of the estimated total tested (Figure S1-C) for each week.

Figure S1-D shows the rate ratio for any case of COVID-19. This was obtained directly from the case-control study using the same definition as in the main manuscript via the same analysis - conditional logisitic regression. Unlike the main analysis, cases who did not have a positive test and their matched controls (i.e. those defined solely on the basis of a hospital discharge diagnostic code or death) were excluded from the comparison, for comparability with the other data shown in the panel plot (Figure S1) which relates to positive tests. Consistent with the fact that this was a working-age population only 595 cases were thus excluded.

Figure S1-E shows a formal comparison of the proportion testing positive of all those tested in a given week, based only on the case-control dataset. This comparison was conducted entirely within the case-control dataset and does not rely the estimated test rates based on the teacher (or healthcare worker) denominator datasets described above. As such it is also possible to include a comparison of household members of teachers and healthcare workers. This comparison was implemented as follows.

First, we restricted the case-control dataset to cases and controls who had been tested. The original stratification (by age in single years, sex and GP practice) would have led to too few strata with at least one case or control tested contemporaneously, therefore we created new strata based on a combination of week of testing and health board area of residence (there are 14 geographic health boards in Scotland). Effect estimates for teachers and healthcare workers were then obtained by fitting a generalised linear model with a logit link and binomial likelihood conditioning on these new stratifying variables (this has previously been shown to be mathematically identical to fitting a conditional logistic regression model although it is less computationally efficient in some circumstances) as well as on the other potential confounders and matching variables included in the main analysis. Figure S1-E shows the results of fitting this model in rolling 3-week periods from March to January 2021.

Healthcare workers were much more likely to be tested for SARS-CoV-2, while the number of teachers tested each week for SARS-CoV-2 was similar to the numbers of individuals in the general population except in late August and late December when there were short spikes in testing among teachers (Figure S1-A). Relative to the general population, the proportion of teachers testing positive (of those tested) increased more rapidly in September before gradually declining towards (but not reaching) the population average in the period from mid-September until the end of follow-up (Figures S1-C and S1-E). Household members of teachers were more similar to the general population (Figure S1-E).

The proportion testing positive in healthcare workers was less than that of the general population. This is paradoxical given the higher rates of severe COVID-19 and hospitalisation with COVID-19 in this group. The finding is too early to be explained by vaccination and the most plausible explanation is that the pattern of testing is different among healthcare workers, with a higher proportion of tests being screening tests than the other groups. We do not know the extent to which different patterns in testing account for differences observed in the proportion testing positive for teachers, household members of teachers, or household members of healthcare workers.

Figure S1 Testing, test positives and test positives as a proportion of those tested over time

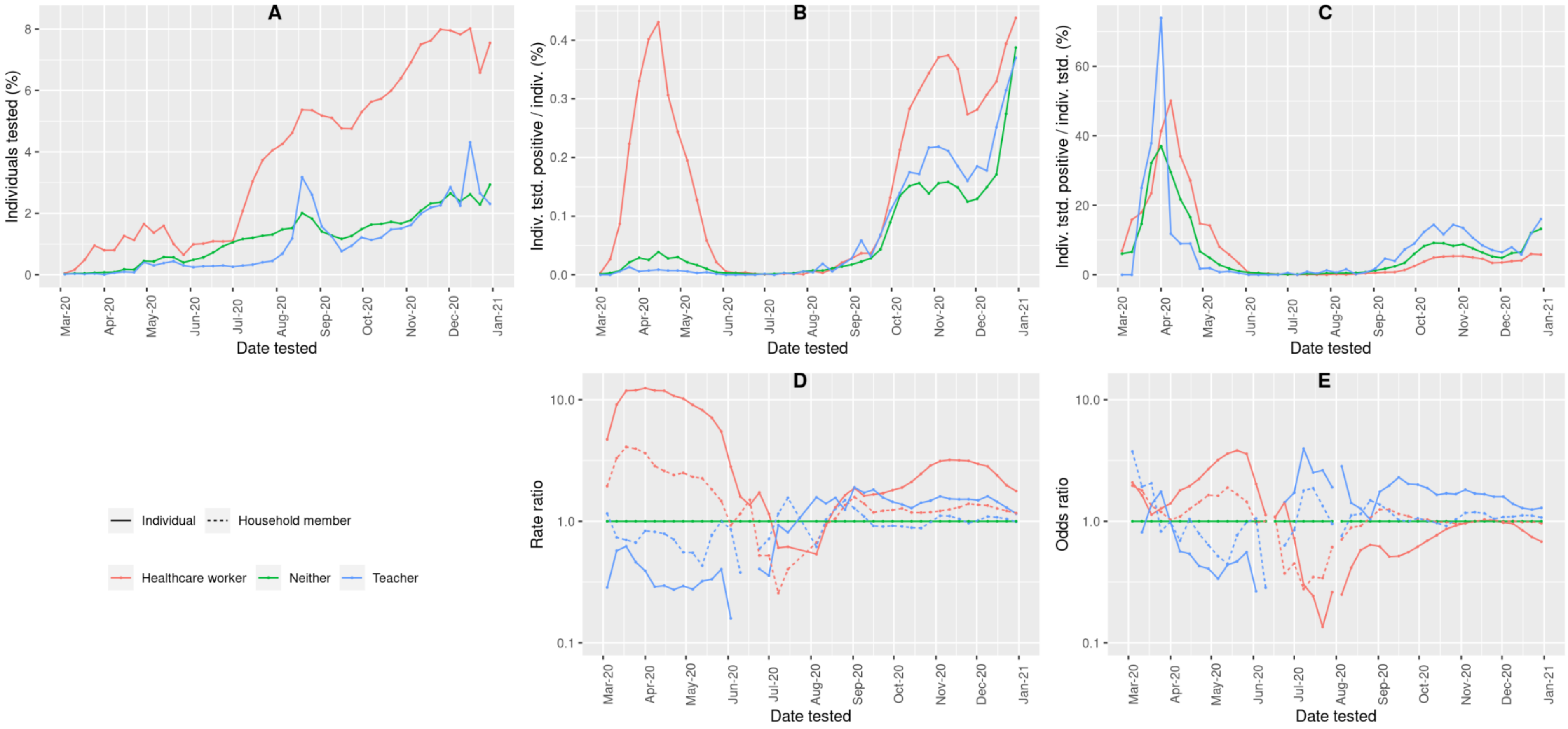


See text above for description of figure.

### Counts of events

Table S1 - Counts of events by age, sex, time period and number of individuals in each group. Counts are mutually exclusive

| Sex | Time period | Age group (years) | Group | Controls | Cases only | Hospitalised not severe | Severe |
|---|---|---|---|---|---|---|---|
| Men | 1 - pre closure | 21 - 30 | Healthcare worker | 17 | 32 | 1 | 0 |
| Men | 1 - pre closure | 21 - 30 | Household member of healthcare worker | 55 | 13 | 2 | 0 |
| Men | 1 - pre closure | 21 - 30 | Household member of teacher | 22 | 4 | 0 | 0 |
| Men | 1 - pre closure | 21 - 30 | Neither | 975 | 31 | 16 | 5 |
| Men | 1 - pre closure | 21 - 30 | Teacher | 9 | 1 | 1 | 0 |
| Men | 2 - closed (before phase 3 easing) | 21 - 30 | Healthcare worker | 58 | 89 | 1 | 1 |
| Men | 2 - closed (before phase 3 easing) | 21 - 30 | Household member of healthcare worker | 166 | 28 | 5 | 1 |
| Men | 2 - closed (before phase 3 easing) | 21 - 30 | Household member of teacher | 104 | 5 | 1 | 0 |
| Men | 2 - closed (before phase 3 easing) | 21 - 30 | Neither | 3931 | 271 | 21 | 6 |
| Men | 2 - closed (before phase 3 easing) | 21 - 30 | Teacher | 25 | 1 | 0 | 0 |
| Men | 2 - closed (phase 3 easing) | 21 - 30 | Healthcare worker | 36 | 2 | 0 | 0 |
| Men | 2 - closed (phase 3 easing) | 21 - 30 | Household member of healthcare worker | 139 | 15 | 0 | 0 |
| Men | 2 - closed (phase 3 easing) | 21 - 30 | Household member of teacher | 93 | 10 | 0 | 0 |
| Men | 2 - closed (phase 3 easing) | 21 - 30 | Neither | 3081 | 298 | 4 | 1 |
| Men | 2 - closed (phase 3 easing) | 21 - 30 | Teacher | 29 | 4 | 0 | 0 |
| Men | 3 - reopened | 21 - 30 | Healthcare worker | 585 | 173 | 6 | 1 |
| Men | 3 - reopened | 21 - 30 | Household member of healthcare worker | 2636 | 338 | 2 | 1 |
| Men | 3 - reopened | 21 - 30 | Household member of teacher | 1948 | 237 | 2 | 0 |
| Men | 3 - reopened | 21 - 30 | Neither | 67373 | 6261 | 94 | 12 |
| Men | 3 - reopened | 21 - 30 | Teacher | 446 | 85 | 1 | 0 |
| Women | 1 - pre closure | 21 - 30 | Healthcare worker | 83 | 67 | 4 | 0 |
| Women | 1 - pre closure | 21 - 30 | Household member of healthcare worker | 37 | 9 | 1 | 0 |
| Women | 1 - pre closure | 21 - 30 | Household member of teacher | 21 | 1 | 0 | 0 |
| Women | 1 - pre closure | 21 - 30 | Neither | 1307 | 37 | 29 | 2 |
| Women | 1 - pre closure | 21 - 30 | Teacher | 53 | 0 | 2 | 0 |
| Women | 2 - closed (before phase 3 easing) | 21 - 30 | Healthcare worker | 527 | 428 | 13 | 1 |
| Women | 2 - closed (before phase 3 easing) | 21 - 30 | Household member of healthcare worker | 308 | 32 | 2 | 0 |
| Women | 2 - closed (before phase 3 easing) | 21 - 30 | Household member of teacher | 181 | 9 | 0 | 0 |

| Sex | Time period | Age group (years) | Group | Controls | Cases only | Hospitalised not severe | Severe |
|---|---|---|---|---|---|---|---|
| Women | 2 - closed (before phase 3 easing) | 21 - 30 | Neither | 10917 | 692 | 47 | 8 |
| Women | 2 - closed (before phase 3 easing) | 21 - 30 | Teacher | 329 | 6 | 0 | 0 |
| Women | 2 - closed (phase 3 easing) | 21 - 30 | Healthcare worker | 102 | 16 | 1 | 0 |
| Women | 2 - closed (phase 3 easing) | 21 - 30 | Household member of healthcare worker | 68 | 8 | 0 | 0 |
| Women | 2 - closed (phase 3 easing) | 21 - 30 | Household member of teacher | 42 | 6 | 0 | 0 |
| Women | 2 - closed (phase 3 easing) | 21 - 30 | Neither | 2368 | 225 | 4 | 0 |
| Women | 2 - closed (phase 3 easing) | 21 - 30 | Teacher | 55 | 6 | 0 | 0 |
| Women | 3 - reopened | 21 - 30 | Healthcare worker | 3397 | 800 | 13 | 1 |
| Women | 3 - reopened | 21 - 30 | Household member of healthcare worker | 2020 | 285 | 2 | 1 |
| Women | 3 - reopened | 21 - 30 | Household member of teacher | 1715 | 174 | 3 | 0 |
| Women | 3 - reopened | 21 - 30 | Neither | 89138 | 8040 | 172 | 19 |
| Women | 3 - reopened | 21 - 30 | Teacher | 2693 | 360 | 6 | 0 |
| Men | 1 - pre closure | 31 - 40 | Healthcare worker | 21 | 28 | 3 | 2 |
| Men | 1 - pre closure | 31 - 40 | Household member of healthcare worker | 57 | 26 | 2 | 0 |
| Men | 1 - pre closure | 31 - 40 | Household member of teacher | 54 | 2 | 1 | 0 |
| Men | 1 - pre closure | 31 - 40 | Neither | 1528 | 47 | 38 | 15 |
| Men | 1 - pre closure | 31 - 40 | Teacher | 13 | 0 | 1 | 0 |
| Men | 2 - closed (before phase 3 easing) | 31 - 40 | Healthcare worker | 65 | 100 | 5 | 0 |
| Men | 2 - closed (before phase 3 easing) | 31 - 40 | Household member of healthcare worker | 160 | 32 | 8 | 1 |
| Men | 2 - closed (before phase 3 easing) | 31 - 40 | Household member of teacher | 138 | 6 | 0 | 0 |
| Men | 2 - closed (before phase 3 easing) | 31 - 40 | Neither | 4577 | 292 | 33 | 16 |
| Men | 2 - closed (before phase 3 easing) | 31 - 40 | Teacher | 55 | 2 | 0 | 0 |
| Men | 2 - closed (phase 3 easing) | 31 - 40 | Healthcare worker | 26 | 3 | 0 | 0 |
| Men | 2 - closed (phase 3 easing) | 31 - 40 | Household member of healthcare worker | 62 | 5 | 0 | 0 |
| Men | 2 - closed (phase 3 easing) | 31 - 40 | Household member of teacher | 43 | 4 | 0 | 0 |
| Men | 2 - closed (phase 3 easing) | 31 - 40 | Neither | 1704 | 167 | 2 | 0 |
| Men | 2 - closed (phase 3 easing) | 31 - 40 | Teacher | 21 | 0 | 0 | 0 |
| Men | 3 - reopened | 31 - 40 | Healthcare worker | 783 | 199 | 2 | 3 |
| Men | 3 - reopened | 31 - 40 | Household member of healthcare worker | 2076 | 292 | 5 | 1 |
| Men | 3 - reopened | 31 - 40 | Household member of teacher | 1938 | 228 | 3 | 0 |
| Men | 3 - reopened | 31 - 40 | Neither | 62814 | 5726 | 157 | 43 |

| Sex | Time period | Age group (years) | Group | Controls | Cases only | Hospitalised not severe | Severe |
|---|---|---|---|---|---|---|---|
| Men | 3 - reopened | 31 - 40 | Teacher | 682 | 127 | 2 | 0 |
| Women | 1 - pre closure | 31 - 40 | Healthcare worker | 122 | 89 | 6 | 0 |
| Women | 1 - pre closure | 31 - 40 | Household member of healthcare worker | 13 | 3 | 0 | 0 |
| Women | 1 - pre closure | 31 - 40 | Household member of teacher | 18 | 2 | 0 | 0 |
| Women | 1 - pre closure | 31 - 40 | Neither | 1828 | 60 | 38 | 9 |
| Women | 1 - pre closure | 31 - 40 | Teacher | 104 | 1 | 1 | 1 |
| Women | 2 - closed (before phase 3 easing) | 31 - 40 | Healthcare worker | 683 | 395 | 15 | 2 |
| Women | 2 - closed (before phase 3 easing) | 31 - 40 | Household member of healthcare worker | 130 | 11 | 1 | 0 |
| Women | 2 - closed (before phase 3 easing) | 31 - 40 | Household member of teacher | 89 | 2 | 0 | 0 |
| Women | 2 - closed (before phase 3 easing) | 31 - 40 | Neither | 10875 | 708 | 75 | 13 |
| Women | 2 - closed (before phase 3 easing) | 31 - 40 | Teacher | 445 | 9 | 2 | 0 |
| Women | 2 - closed (phase 3 easing) | 31 - 40 | Healthcare worker | 91 | 4 | 1 | 0 |
| Women | 2 - closed (phase 3 easing) | 31 - 40 | Household member of healthcare worker | 8 | 1 | 0 | 0 |
| Women | 2 - closed (phase 3 easing) | 31 - 40 | Household member of teacher | 13 | 0 | 0 | 0 |
| Women | 2 - closed (phase 3 easing) | 31 - 40 | Neither | 1343 | 128 | 6 | 1 |
| Women | 2 - closed (phase 3 easing) | 31 - 40 | Teacher | 55 | 9 | 0 | 0 |
| Women | 3 - reopened | 31 - 40 | Healthcare worker | 4313 | 916 | 25 | 2 |
| Women | 3 - reopened | 31 - 40 | Household member of healthcare worker | 711 | 82 | 2 | 1 |
| Women | 3 - reopened | 31 - 40 | Household member of teacher | 761 | 57 | 0 | 0 |
| Women | 3 - reopened | 31 - 40 | Neither | 83842 | 7532 | 244 | 37 |
| Women | 3 - reopened | 31 - 40 | Teacher | 3577 | 416 | 15 | 1 |
| Men | 1 - pre closure | 41 - 50 | Healthcare worker | 50 | 54 | 10 | 2 |
| Men | 1 - pre closure | 41 - 50 | Household member of healthcare worker | 85 | 21 | 7 | 1 |
| Men | 1 - pre closure | 41 - 50 | Household member of teacher | 65 | 1 | 0 | 0 |
| Men | 1 - pre closure | 41 - 50 | Neither | 2418 | 49 | 75 | 45 |
| Men | 1 - pre closure | 41 - 50 | Teacher | 25 | 1 | 0 | 0 |
| Men | 2 - closed (before phase 3 easing) | 41 - 50 | Healthcare worker | 105 | 130 | 14 | 2 |
| Men | 2 - closed (before phase 3 easing) | 41 - 50 | Household member of healthcare worker | 231 | 35 | 10 | 1 |
| Men | 2 - closed (before phase 3 easing) | 41 - 50 | Household member of teacher | 158 | 4 | 1 | 0 |
| Men | 2 - closed (before phase 3 easing) | 41 - 50 | Neither | 5679 | 271 | 99 | 44 |
| Men | 2 - closed (before phase 3 easing) | 41 - 50 | Teacher | 42 | 4 | 0 | 0 |

| Sex | Time period | Age group (years) | Group | Controls | Cases only | Hospitalised not severe | Severe |
|---|---|---|---|---|---|---|---|
| Men | 2 - closed (phase 3 easing) | 41 - 50 | Healthcare worker | 30 | 5 | 1 | 0 |
| Men | 2 - closed (phase 3 easing) | 41 - 50 | Household member of healthcare worker | 40 | 4 | 0 | 0 |
| Men | 2 - closed (phase 3 easing) | 41 - 50 | Household member of teacher | 36 | 5 | 0 | 0 |
| Men | 2 - closed (phase 3 easing) | 41 - 50 | Neither | 1416 | 119 | 9 | 4 |
| Men | 2 - closed (phase 3 easing) | 41 - 50 | Teacher | 16 | 4 | 0 | 0 |
| Men | 3 - reopened | 41 - 50 | Healthcare worker | 989 | 221 | 8 | 1 |
| Men | 3 - reopened | 41 - 50 | Household member of healthcare worker | 1897 | 297 | 12 | 2 |
| Men | 3 - reopened | 41 - 50 | Household member of teacher | 1652 | 199 | 9 | 2 |
| Men | 3 - reopened | 41 - 50 | Neither | 60144 | 5256 | 285 | 89 |
| Men | 3 - reopened | 41 - 50 | Teacher | 571 | 98 | 6 | 1 |
| Women | 1 - pre closure | 41 - 50 | Healthcare worker | 206 | 113 | 14 | 6 |
| Women | 1 - pre closure | 41 - 50 | Household member of healthcare worker | 32 | 7 | 2 | 0 |
| Women | 1 - pre closure | 41 - 50 | Household member of teacher | 23 | 0 | 0 | 2 |
| Women | 1 - pre closure | 41 - 50 | Neither | 2533 | 78 | 52 | 16 |
| Women | 1 - pre closure | 41 - 50 | Teacher | 118 | 1 | 2 | 0 |
| Women | 2 - closed (before phase 3 easing) | 41 - 50 | Healthcare worker | 859 | 420 | 34 | 3 |
| Women | 2 - closed (before phase 3 easing) | 41 - 50 | Household member of healthcare worker | 138 | 26 | 1 | 2 |
| Women | 2 - closed (before phase 3 easing) | 41 - 50 | Household member of teacher | 125 | 3 | 0 | 0 |
| Women | 2 - closed (before phase 3 easing) | 41 - 50 | Neither | 12917 | 802 | 124 | 36 |
| Women | 2 - closed (before phase 3 easing) | 41 - 50 | Teacher | 483 | 4 | 1 | 1 |
| Women | 2 - closed (phase 3 easing) | 41 - 50 | Healthcare worker | 68 | 7 | 0 | 0 |
| Women | 2 - closed (phase 3 easing) | 41 - 50 | Household member of healthcare worker | 6 | 0 | 0 | 0 |
| Women | 2 - closed (phase 3 easing) | 41 - 50 | Household member of teacher | 9 | 1 | 0 | 0 |
| Women | 2 - closed (phase 3 easing) | 41 - 50 | Neither | 1317 | 126 | 5 | 1 |
| Women | 2 - closed (phase 3 easing) | 41 - 50 | Teacher | 51 | 5 | 0 | 0 |
| Women | 3 - reopened | 41 - 50 | Healthcare worker | 4632 | 890 | 34 | 5 |
| Women | 3 - reopened | 41 - 50 | Household member of healthcare worker | 763 | 84 | 4 | 1 |
| Women | 3 - reopened | 41 - 50 | Household member of teacher | 658 | 67 | 3 | 0 |
| Women | 3 - reopened | 41 - 50 | Neither | 78980 | 7006 | 303 | 73 |
| Women | 3 - reopened | 41 - 50 | Teacher | 2817 | 330 | 11 | 0 |
| Men | 1 - pre closure | 51 - 65 | Healthcare worker | 129 | 64 | 9 | 2 |

| Sex | Time period | Age group (years) | Group | Controls | Cases only | Hospitalised not severe | Severe |
|---|---|---|---|---|---|---|---|
| Men | 1 - pre closure | 51 - 65 | Household member of healthcare worker | 233 | 35 | 17 | 9 |
| Men | 1 - pre closure | 51 - 65 | Household member of teacher | 153 | 2 | 3 | 3 |
| Men | 1 - pre closure | 51 - 65 | Neither | 5802 | 103 | 211 | 176 |
| Men | 1 - pre closure | 51 - 65 | Teacher | 40 | 0 | 1 | 1 |
| Men | 2 - closed (before phase 3 easing) | 51 - 65 | Healthcare worker | 229 | 142 | 20 | 7 |
| Men | 2 - closed (before phase 3 easing) | 51 - 65 | Household member of healthcare worker | 460 | 66 | 20 | 15 |
| Men | 2 - closed (before phase 3 easing) | 51 - 65 | Household member of teacher | 278 | 16 | 5 | 5 |
| Men | 2 - closed (before phase 3 easing) | 51 - 65 | Neither | 12317 | 477 | 303 | 179 |
| Men | 2 - closed (before phase 3 easing) | 51 - 65 | Teacher | 70 | 3 | 0 | 1 |
| Men | 2 - closed (phase 3 easing) | 51 - 65 | Healthcare worker | 27 | 1 | 0 | 0 |
| Men | 2 - closed (phase 3 easing) | 51 - 65 | Household member of healthcare worker | 57 | 7 | 0 | 1 |
| Men | 2 - closed (phase 3 easing) | 51 - 65 | Household member of teacher | 35 | 5 | 1 | 0 |
| Men | 2 - closed (phase 3 easing) | 51 - 65 | Neither | 1422 | 123 | 7 | 7 |
| Men | 2 - closed (phase 3 easing) | 51 - 65 | Teacher | 6 | 2 | 0 | 0 |
| Men | 3 - reopened | 51 - 65 | Healthcare worker | 1569 | 349 | 28 | 6 |
| Men | 3 - reopened | 51 - 65 | Household member of healthcare worker | 3076 | 455 | 31 | 15 |
| Men | 3 - reopened | 51 - 65 | Household member of teacher | 2271 | 271 | 21 | 8 |
| Men | 3 - reopened | 51 - 65 | Neither | 94451 | 7638 | 809 | 435 |
| Men | 3 - reopened | 51 - 65 | Teacher | 610 | 80 | 2 | 0 |
| Women | 1 - pre closure | 51 - 65 | Healthcare worker | 346 | 149 | 11 | 8 |
| Women | 1 - pre closure | 51 - 65 | Household member of healthcare worker | 71 | 10 | 0 | 1 |
| Women | 1 - pre closure | 51 - 65 | Household member of teacher | 69 | 0 | 3 | 0 |
| Women | 1 - pre closure | 51 - 65 | Neither | 4927 | 111 | 173 | 82 |
| Women | 1 - pre closure | 51 - 65 | Teacher | 124 | 2 | 1 | 2 |
| Women | 2 - closed (before phase 3 easing) | 51 - 65 | Healthcare worker | 1237 | 533 | 52 | 10 |
| Women | 2 - closed (before phase 3 easing) | 51 - 65 | Household member of healthcare worker | 282 | 43 | 7 | 3 |
| Women | 2 - closed (before phase 3 easing) | 51 - 65 | Household member of teacher | 265 | 5 | 5 | 1 |
| Women | 2 - closed (before phase 3 easing) | 51 - 65 | Neither | 21077 | 1182 | 323 | 117 |
| Women | 2 - closed (before phase 3 easing) | 51 - 65 | Teacher | 458 | 3 | 0 | 0 |
| Women | 2 - closed (phase 3 easing) | 51 - 65 | Healthcare worker | 126 | 14 | 0 | 0 |
| Women | 2 - closed (phase 3 easing) | 51 - 65 | Household member of healthcare worker | 28 | 0 | 0 | 0 |

| Sex | Time period | Age group (years) | Group | Controls | Cases only | Hospitalised not severe | Severe |
|---|---|---|---|---|---|---|---|
| Women | 2 - closed (phase 3 easing) | 51 - 65 | Household member of teacher | 30 | 4 | 0 | 0 |
| Women | 2 - closed (phase 3 easing) | 51 - 65 | Neither | 1752 | 159 | 10 | 4 |
| Women | 2 - closed (phase 3 easing) | 51 - 65 | Teacher | 35 | 6 | 0 | 0 |
| Women | 3 - reopened | 51 - 65 | Healthcare worker | 5896 | 1149 | 57 | 19 |
| Women | 3 - reopened | 51 - 65 | Household member of healthcare worker | 1297 | 173 | 18 | 6 |
| Women | 3 - reopened | 51 - 65 | Household member of teacher | 1462 | 170 | 10 | 1 |
| Women | 3 - reopened | 51 - 65 | Neither | 111409 | 9341 | 754 | 239 |
| Women | 3 - reopened | 51 - 65 | Teacher | 2257 | 245 | 8 | 2 |

### Effect measure estimates

The following tables display the same effect estimates as shown Tables 2 and 3 of the main manuscript but broken down into age and sex strata (Table S2a and S2b), additional time periods (Tables S3a and S3b) or age and sex strata and additional time periods (Tables S4a and S4b). Please see footnotes of Tables 2 and 3 of the main manuscript for additional details.

Table S2a - Rate ratios for any case and hospitalisation with COVID-19 for teachers, healthcare workers and members of their households, entire time period, stratified by age and sex

| Adjustment | Time period | Sex | Age group (years) | Outcome | Neither | Healthcare worker | Household member of healthcare worker | Household member of teacher | Teacher |
|---|---|---|---|---|---|---|---|---|---|
| Unadjusted | Full period | Men | 21 - 30 | Any case | 1 | 4.89 (4.25-5.62) | 1.45 (1.31-1.61) | 1.25 (1.09-1.42) | 1.89 (1.51-2.36) |
| Adjusted | Full period | Men | 21 - 30 | Any case | 1 | 4.95 (4.29-5.72) | 1.27 (1.14-1.41) | 1.12 (0.98-1.27) | 1.87 (1.49-2.34) |
| Unadjusted | Full period | Men | 21 - 30 | Hospitalisation | 1 | 7.87 (3.28-18.85) | 1.79 (0.92-3.51) | 0.71 (0.22-2.31) | 1.96 (0.42-9.09) |
| Adjusted | Full period | Men | 21 - 30 | Hospitalisation | 1 | 9.44 (3.66-24.34) | 1.86 (0.92-3.76) | 0.57 (0.15-2.18) | 1.68 (0.32-8.75) |
| Unadjusted | Full period | Men | 31 - 40 | Any case | 1 | 4.24 (3.73-4.82) | 1.68 (1.51-1.88) | 1.21 (1.06-1.38) | 1.78 (1.48-2.15) |
| Adjusted | Full period | Men | 31 - 40 | Any case | 1 | 4.28 (3.74-4.89) | 1.51 (1.35-1.70) | 1.11 (0.97-1.27) | 1.78 (1.47-2.15) |
| Unadjusted | Full period | Men | 31 - 40 | Hospitalisation | 1 | 4.02 (2.12-7.61) | 2.07 (1.20-3.60) | 0.48 (0.18-1.33) | 1.13 (0.34-3.76) |
| Adjusted | Full period | Men | 31 - 40 | Hospitalisation | 1 | 4.94 (2.44-10.01) | 1.99 (1.12-3.53) | 0.49 (0.18-1.38) | 1.03 (0.30-3.54) |
| Unadjusted | Full period | Men | 41 - 50 | Any case | 1 | 4.23 (3.78-4.74) | 1.90 (1.70-2.12) | 1.24 (1.08-1.43) | 1.83 (1.50-2.24) |
| Adjusted | Full period | Men | 41 - 50 | Any case | 1 | 4.23 (3.76-4.77) | 1.51 (1.35-1.69) | 1.02 (0.88-1.18) | 1.86 (1.51-2.29) |
| Unadjusted | Full period | Men | 41 - 50 | Hospitalisation | 1 | 3.28 (2.26-4.78) | 1.57 (1.08-2.27) | 0.92 (0.54-1.58) | 1.24 (0.56-2.73) |
| Adjusted | Full period | Men | 41 - 50 | Hospitalisation | 1 | 3.71 (2.51-5.49) | 1.35 (0.91-2.01) | 1.04 (0.59-1.81) | 1.33 (0.58-3.05) |
| Unadjusted | Full period | Men | 51 - 65 | Any case | 1 | 3.55 (3.23-3.90) | 1.92 (1.76-2.08) | 1.30 (1.17-1.46) | 1.30 (1.04-1.62) |
| Adjusted | Full period | Men | 51 - 65 | Any case | 1 | 3.46 (3.13-3.82) | 1.47 (1.35-1.60) | 0.97 (0.87-1.10) | 1.19 (0.94-1.49) |
| Unadjusted | Full period | Men | 51 - 65 | Hospitalisation | 1 | 1.96 (1.51-2.53) | 1.54 (1.26-1.89) | 0.96 (0.70-1.32) | 0.37 (0.15-0.92) |

| Adjustment | Time period | Sex | Age group (years) | Outcome | Neither | Healthcare worker | Household member of healthcare worker | Household member of teacher | Teacher |
|---|---|---|---|---|---|---|---|---|---|
| Adjusted | Full period | Men | 51 - 65 | Hospitalisation | 1 | 1.96 (1.50-2.56) | 1.34 (1.09-1.67) | 0.86 (0.62-1.19) | 0.31 (0.12-0.78) |
| Unadjusted | Full period | Women | 21 - 30 | Any case | 1 | 3.75 (3.51-4.01) | 1.53 (1.36-1.71) | 1.02 (0.88-1.18) | 1.36 (1.22-1.52) |
| Adjusted | Full period | Women | 21 - 30 | Any case | 1 | 3.85 (3.60-4.12) | 1.33 (1.18-1.49) | 0.89 (0.77-1.03) | 1.39 (1.25-1.56) |
| Unadjusted | Full period | Women | 21 - 30 | Hospitalisation | 1 | 2.93 (1.94-4.42) | 0.99 (0.45-2.20) | 0.54 (0.17-1.75) | 0.86 (0.41-1.80) |
| Adjusted | Full period | Women | 21 - 30 | Hospitalisation | 1 | 3.08 (2.00-4.76) | 0.97 (0.42-2.23) | 0.73 (0.21-2.51) | 0.93 (0.43-2.02) |
| Unadjusted | Full period | Women | 31 - 40 | Any case | 1 | 3.16 (2.97-3.37) | 1.24 (1.02-1.52) | 0.84 (0.67-1.06) | 1.22 (1.11-1.35) |
| Adjusted | Full period | Women | 31 - 40 | Any case | 1 | 3.29 (3.08-3.51) | 1.09 (0.89-1.34) | 0.76 (0.61-0.96) | 1.31 (1.19-1.45) |
| Unadjusted | Full period | Women | 31 - 40 | Hospitalisation | 1 | 2.33 (1.69-3.22) | 1.16 (0.46-2.95) | 0.21 (0.03-1.51) | 1.14 (0.71-1.84) |
| Adjusted | Full period | Women | 31 - 40 | Hospitalisation | 1 | 2.58 (1.83-3.64) | 0.84 (0.31-2.26) | 0.17 (0.02-1.36) | 1.21 (0.74-1.99) |
| Unadjusted | Full period | Women | 41 - 50 | Any case | 1 | 2.98 (2.80-3.17) | 1.53 (1.28-1.83) | 1.04 (0.84-1.28) | 1.14 (1.02-1.28) |
| Adjusted | Full period | Women | 41 - 50 | Any case | 1 | 3.00 (2.82-3.20) | 1.25 (1.04-1.51) | 0.85 (0.68-1.05) | 1.13 (1.00-1.26) |
| Unadjusted | Full period | Women | 41 - 50 | Hospitalisation | 1 | 2.61 (2.05-3.33) | 1.60 (0.84-3.02) | 1.06 (0.48-2.34) | 0.74 (0.43-1.26) |
| Adjusted | Full period | Women | 41 - 50 | Hospitalisation | 1 | 2.93 (2.26-3.80) | 1.37 (0.69-2.74) | 1.15 (0.50-2.62) | 0.86 (0.49-1.49) |
| Unadjusted | Full period | Women | 51 - 65 | Any case | 1 | 2.98 (2.82-3.14) | 1.77 (1.56-2.01) | 1.16 (1.01-1.33) | 1.04 (0.92-1.18) |
| Adjusted | Full period | Women | 51 - 65 | Any case | 1 | 2.98 (2.82-3.15) | 1.36 (1.19-1.55) | 0.87 (0.76-1.00) | 1.01 (0.89-1.16) |
| Unadjusted | Full period | Women | 51 - 65 | Hospitalisation | 1 | 1.76 (1.47-2.10) | 1.70 (1.18-2.46) | 0.90 (0.57-1.42) | 0.42 (0.24-0.73) |
| Adjusted | Full period | Women | 51 - 65 | Hospitalisation | 1 | 1.92 (1.60-2.32) | 1.34 (0.90-1.99) | 0.77 (0.48-1.25) | 0.51 (0.29-0.90) |

Table S2b Rate ratios for any case and hospitalisation with COVID-19 for teachers by sector, entire time period, stratified by age and sex

| **Adjustment** | **Time period** | **Sex** | **Age group (years)** | **Outcome** | **Neither** | **Nursery/Primary or Nursery** | **Primary** | **Secondary** | **Teacher in other sector** |
|---|---|---|---|---|---|---|---|---|---|
| Unadjusted | Full period | Men | 21 - 30 | Any case | 1 | 2.13 (1.22-3.72) | 1.97 (1.15-3.38) | 1.71 (1.27-2.30) | 2.78 (1.37-5.61) |
| Adjusted | Full period | Men | 21 - 30 | Any case | 1 | 1.96 (1.11-3.47) | 2.07 (1.19-3.60) | 1.69 (1.25-2.28) | 2.76 (1.34-5.65) |
| Unadjusted | Full period | Men | 21 - 30 | Hospitalisation | 1 | - | - | 1.15 (0.14-9.36) | - |
| Adjusted | Full period | Men | 21 - 30 | Hospitalisation | 1 | - | - | 0.78 (0.09-7.05) | - |
| Unadjusted | Full period | Men | 31 - 40 | Any case | 1 | 1.52 (0.91-2.54) | 1.86 (1.14-3.03) | 1.83 (1.43-2.32) | 1.81 (1.08-3.05) |
| Adjusted | Full period | Men | 31 - 40 | Any case | 1 | 1.53 (0.91-2.57) | 1.77 (1.07-2.93) | 1.82 (1.42-2.33) | 1.90 (1.11-3.24) |
| Unadjusted | Full period | Men | 31 - 40 | Hospitalisation | 1 | 1.80 (0.22-15.01) | - | 1.13 (0.26-4.91) | - |
| Adjusted | Full period | Men | 31 - 40 | Hospitalisation | 1 | 1.78 (0.20-15.64) | - | 0.99 (0.22-4.59) | - |
| Unadjusted | Full period | Men | 41 - 50 | Any case | 1 | 1.43 (0.74-2.78) | 3.40 (2.02-5.73) | 1.50 (1.14-1.97) | 2.48 (1.63-3.79) |
| Adjusted | Full period | Men | 41 - 50 | Any case | 1 | 1.52 (0.77-3.00) | 3.00 (1.73-5.19) | 1.55 (1.16-2.06) | 2.53 (1.63-3.93) |
| Unadjusted | Full period | Men | 41 - 50 | Hospitalisation | 1 | 1.24 (0.16-9.95) | - | 1.17 (0.41-3.34) | 2.00 (0.44-9.16) |
| Adjusted | Full period | Men | 41 - 50 | Hospitalisation | 1 | 1.16 (0.14-9.90) | - | 1.24 (0.41-3.74) | 2.64 (0.56-12.39) |
| Unadjusted | Full period | Men | 51 - 65 | Any case | 1 | 1.98 (0.88-4.46) | 0.60 (0.19-1.92) | 1.39 (1.06-1.81) | 1.11 (0.67-1.84) |
| Adjusted | Full period | Men | 51 - 65 | Any case | 1 | 1.79 (0.77-4.13) | 0.51 (0.16-1.65) | 1.25 (0.95-1.64) | 1.09 (0.65-1.84) |
| Unadjusted | Full period | Men | 51 - 65 | Hospitalisation | 1 | - | - | 0.48 (0.18-1.32) | 0.29 (0.04-2.16) |
| Adjusted | Full period | Men | 51 - 65 | Hospitalisation | 1 | - | - | 0.41 (0.15-1.16) | 0.21 (0.03-1.73) |

| | | | | | | | | | |
|---|---|---|---|---|---|---|---|---|---|
| Unadjusted | Full period | Women | 21 - 30 | Any case | 1 | 1.23 (0.99-1.54) | 1.58 (1.31-1.89) | 1.30 (1.08-1.56) | 1.24 (0.83-1.87) |
| Adjusted | Full period | Women | 21 - 30 | Any case | 1 | 1.23 (0.99-1.54) | 1.63 (1.36-1.96) | 1.34 (1.11-1.62) | 1.24 (0.82-1.87) |
| Unadjusted | Full period | Women | 21 - 30 | Hospitalisation | 1 | 0.62 (0.08-4.72) | 1.38 (0.53-3.58) | 0.28 (0.04-2.06) | 1.81 (0.22-15.11) |
| Adjusted | Full period | Women | 21 - 30 | Hospitalisation | 1 | 0.73 (0.09-5.69) | 1.57 (0.59-4.18) | 0.25 (0.03-2.01) | 1.81 (0.21-15.88) |
| Unadjusted | Full period | Women | 31 - 40 | Any case | 1 | 1.13 (0.93-1.37) | 1.36 (1.14-1.62) | 1.17 (0.98-1.39) | 1.26 (0.95-1.66) |
| Adjusted | Full period | Women | 31 - 40 | Any case | 1 | 1.23 (1.01-1.50) | 1.49 (1.24-1.78) | 1.23 (1.03-1.47) | 1.32 (0.99-1.75) |
| Unadjusted | Full period | Women | 31 - 40 | Hospitalisation | 1 | 0.68 (0.21-2.19) | 0.59 (0.18-1.90) | 1.69 (0.86-3.32) | 2.00 (0.68-5.86) |
| Adjusted | Full period | Women | 31 - 40 | Hospitalisation | 1 | 0.71 (0.21-2.34) | 0.58 (0.17-1.92) | 1.85 (0.91-3.76) | 2.51 (0.81-7.79) |
| Unadjusted | Full period | Women | 41 - 50 | Any case | 1 | 0.99 (0.79-1.25) | 1.38 (1.12-1.70) | 1.11 (0.91-1.35) | 1.12 (0.85-1.46) |
| Adjusted | Full period | Women | 41 - 50 | Any case | 1 | 0.97 (0.77-1.22) | 1.37 (1.10-1.70) | 1.11 (0.91-1.35) | 1.09 (0.83-1.44) |
| Unadjusted | Full period | Women | 41 - 50 | Hospitalisation | 1 | 0.22 (0.03-1.59) | 0.52 (0.16-1.65) | 0.87 (0.37-2.01) | 1.70 (0.66-4.39) |
| Adjusted | Full period | Women | 41 - 50 | Hospitalisation | 1 | 0.36 (0.05-2.69) | 0.46 (0.13-1.56) | 1.14 (0.48-2.68) | 1.57 (0.59-4.19) |
| Unadjusted | Full period | Women | 51 - 65 | Any case | 1 | 1.08 (0.84-1.38) | 1.45 (1.15-1.81) | 0.83 (0.65-1.06) | 0.85 (0.63-1.16) |
| Adjusted | Full period | Women | 51 - 65 | Any case | 1 | 1.04 (0.81-1.34) | 1.35 (1.07-1.71) | 0.82 (0.63-1.05) | 0.89 (0.65-1.21) |
| Unadjusted | Full period | Women | 51 - 65 | Hospitalisation | 1 | 0.30 (0.07-1.23) | 0.28 (0.07-1.16) | 0.67 (0.31-1.45) | 0.29 (0.07-1.19) |
| Adjusted | Full period | Women | 51 - 65 | Hospitalisation | 1 | 0.33 (0.08-1.36) | 0.32 (0.08-1.31) | 0.88 (0.40-1.92) | 0.40 (0.10-1.65) |

Table S3a - Rate ratios for any case, hospitalisation with COVID-19 and severe COVID-19 for teachers, healthcare workers and members of their households, stratified by time period

| Adjustment | Time period | Outcome | Neither | Healthcare worker | Household member of healthcare worker | Household member of teacher | Teacher |
|---|---|---|---|---|---|---|---|
| Unadjusted | 1 - pre closure | Any case | 1 | 11.12 (9.85-12.55) | 4.15 (3.45-4.99) | 0.82 (0.58-1.16) | 0.57 (0.36-0.92) |
| Adjusted | 1 - pre closure | Any case | 1 | 11.41 (10.07-12.92) | 3.92 (3.25-4.73) | 0.78 (0.55-1.11) | 0.57 (0.36-0.92) |
| Unadjusted | 1 - pre closure | Hospitalisation | 1 | 2.23 (1.72-2.88) | 1.70 (1.22-2.36) | 0.66 (0.38-1.15) | 0.87 (0.49-1.56) |
| Adjusted | 1 - pre closure | Hospitalisation | 1 | 2.32 (1.78-3.02) | 1.49 (1.06-2.10) | 0.67 (0.38-1.17) | 0.95 (0.53-1.70) |
| Unadjusted | 1 - pre closure | Severe | 1 | 1.99 (1.22-3.25) | 1.57 (0.84-2.93) | 0.86 (0.37-2.00) | 0.91 (0.32-2.57) |
| Adjusted | 1 - pre closure | Severe | 1 | 2.11 (1.27-3.51) | 1.49 (0.78-2.85) | 0.81 (0.34-1.92) | 0.95 (0.33-2.71) |
| Unadjusted | 2 - closed (before phase 3 easing) | Any case | 1 | 9.49 (8.93-10.10) | 2.50 (2.22-2.81) | 0.73 (0.59-0.90) | 0.27 (0.19-0.37) |
| Adjusted | 2 - closed (before phase 3 easing) | Any case | 1 | 9.80 (9.20-10.43) | 2.33 (2.07-2.63) | 0.71 (0.58-0.88) | 0.29 (0.21-0.40) |
| Unadjusted | 2 - closed (before phase 3 easing) | Hospitalisation | 1 | 3.47 (2.89-4.15) | 2.32 (1.80-2.98) | 0.67 (0.41-1.09) | 0.21 (0.09-0.51) |
| Adjusted | 2 - closed (before phase 3 easing) | Hospitalisation | 1 | 3.92 (3.25-4.74) | 2.24 (1.72-2.93) | 0.64 (0.39-1.06) | 0.24 (0.10-0.59) |
| Unadjusted | 2 - closed (before phase 3 easing) | Severe | 1 | 2.05 (1.33-3.17) | 2.26 (1.43-3.57) | 0.94 (0.40-2.17) | 0.34 (0.08-1.39) |
| Adjusted | 2 - closed (before phase 3 easing) | Severe | 1 | 2.15 (1.35-3.42) | 2.59 (1.57-4.30) | 0.88 (0.36-2.18) | 0.43 (0.10-1.86) |
| Unadjusted | 2 - closed (phase 3 easing) | Any case | 1 | 1.11 (0.83-1.48) | 1.02 (0.74-1.40) | 1.28 (0.91-1.79) | 1.37 (0.96-1.96) |
| Adjusted | 2 - closed (phase 3 easing) | Any case | 1 | 1.05 (0.79-1.41) | 0.89 (0.65-1.23) | 1.10 (0.78-1.55) | 1.29 (0.90-1.85) |
| Unadjusted | 2 - closed (phase 3 easing) | Hospitalisation | 1 | 1.20 (0.34-4.25) | 0.69 (0.09-5.35) | 0.47 (0.06-3.56) | - |
| Adjusted | 2 - closed (phase 3 easing) | Hospitalisation | 1 | - | - | - | - |
| Unadjusted | 2 - closed (phase 3 easing) | Severe | 1 | - | - | - | - |

| Adjustment | Time period | Outcome | Neither | Healthcare worker | Household member of healthcare worker | Household member of teacher | Teacher |
|---|---|---|---|---|---|---|---|
| Adjusted | 2 - closed (phase 3 easing) | Severe | 1 | - | - | - | - |
| Unadjusted | 3 - reopened | Any case | 1 | 2.41 (2.33-2.49) | 1.54 (1.47-1.61) | 1.22 (1.16-1.29) | 1.41 (1.34-1.49) |
| Adjusted | 3 - reopened | Any case | 1 | 2.41 (2.33-2.49) | 1.25 (1.20-1.31) | 1.01 (0.95-1.06) | 1.42 (1.35-1.49) |
| Unadjusted | 3 - reopened | Hospitalisation | 1 | 1.71 (1.47-2.00) | 1.25 (1.02-1.54) | 1.00 (0.77-1.30) | 0.89 (0.67-1.18) |
| Adjusted | 3 - reopened | Hospitalisation | 1 | 1.82 (1.55-2.14) | 1.04 (0.83-1.29) | 0.97 (0.74-1.27) | 0.97 (0.72-1.29) |
| Unadjusted | 3 - reopened | Severe | 1 | 1.44 (1.02-2.05) | 1.33 (0.90-1.97) | 0.68 (0.38-1.23) | 0.30 (0.11-0.81) |
| Adjusted | 3 - reopened | Severe | 1 | 1.76 (1.22-2.56) | 1.13 (0.74-1.73) | 0.67 (0.36-1.24) | 0.27 (0.09-0.77) |

Table S3b Rate ratios for any case and hospitalisation with COVID-19 for teachers by sector, stratified by time period

| Adjustment | Time period | Outcome | Neither | Nursery/Primary or Nursery | Primary | Secondary | Teacher in other sector |
|---|---|---|---|---|---|---|---|
| Unadjusted | 1 - pre closure | Any case | 1 | 0.12 (0.02-0.89) | 0.71 (0.26-1.97) | 0.71 (0.36-1.41) | 0.73 (0.29-1.87) |
| Adjusted | 1 - pre closure | Any case | 1 | 0.13 (0.02-0.94) | 0.67 (0.24-1.86) | 0.71 (0.36-1.40) | 0.76 (0.30-1.94) |
| Unadjusted | 1 - pre closure | Hospitalisation | 1 | - | 1.46 (0.51-4.16) | 1.03 (0.44-2.39) | 0.91 (0.27-3.05) |
| Adjusted | 1 - pre closure | Hospitalisation | 1 | - | 1.60 (0.55-4.64) | 1.11 (0.47-2.61) | 0.99 (0.29-3.39) |
| Unadjusted | 2 - closed (before phase 3 easing) | Any case | 1 | 0.26 (0.13-0.51) | 0.28 (0.15-0.53) | 0.22 (0.12-0.40) | 0.38 (0.18-0.80) |
| Adjusted | 2 - closed (before phase 3 easing) | Any case | 1 | 0.28 (0.15-0.55) | 0.30 (0.16-0.57) | 0.23 (0.13-0.43) | 0.40 (0.19-0.86) |
| Unadjusted | 2 - closed (before phase 3 easing) | Hospitalisation | 1 | 0.49 (0.12-2.04) | - | 0.22 (0.05-0.87) | 0.28 (0.04-2.05) |
| Adjusted | 2 - closed (before phase 3 easing) | Hospitalisation | 1 | 0.55 (0.13-2.35) | - | 0.25 (0.06-1.01) | 0.33 (0.04-2.41) |
| Unadjusted | 2 - closed (phase 3 easing) | Any case | 1 | 0.99 (0.45-2.17) | 1.58 (0.78-3.20) | 0.92 (0.47-1.84) | 3.03 (1.52-6.03) |
| Adjusted | 2 - closed (phase 3 easing) | Any case | 1 | 0.94 (0.43-2.07) | 1.64 (0.80-3.36) | 0.85 (0.42-1.70) | 2.54 (1.26-5.14) |
| Unadjusted | 2 - closed (phase 3 easing) | Hospitalisation | 1 | - | - | - | - |
| Adjusted | 2 - closed (phase 3 easing) | Hospitalisation | 1 | - | - | - | - |
| Unadjusted | 3 - reopened | Any case | 1 | 1.30 (1.17-1.44) | 1.64 (1.49-1.80) | 1.38 (1.28-1.50) | 1.30 (1.14-1.49) |
| Adjusted | 3 - reopened | Any case | 1 | 1.30 (1.17-1.45) | 1.66 (1.50-1.83) | 1.39 (1.27-1.51) | 1.31 (1.14-1.51) |
| Unadjusted | 3 - reopened | Hospitalisation | 1 | 0.57 (0.27-1.22) | 0.73 (0.38-1.38) | 1.06 (0.71-1.58) | 1.08 (0.58-2.01) |
| Adjusted | 3 - reopened | Hospitalisation | 1 | 0.61 (0.28-1.33) | 0.72 (0.37-1.40) | 1.22 (0.81-1.86) | 1.17 (0.60-2.27) |

Table S4a - Rate ratios for any case and hospitalisation with COVID-19 for teachers, healthcare workers and members of their households, stratified by time period, age and sex

| Adjustment | Time period | Sex | Age group (years) | Outcome | Neither | Healthcare worker | Household member of healthcare worker | Household member of teacher | Teacher |
|---|---|---|---|---|---|---|---|---|---|
| Unadjusted | 1 - pre closure | Men | 21 - 30 | Hospitalisation | 1 | 2.21 (0.21-22.98) | 2.99 (0.54-16.44) | - | 5.51 (0.49-61.45) |
| Unadjusted | 1 - pre closure | Men | 31 - 40 | Hospitalisation | 1 | 6.57 (2.07-20.90) | 1.14 (0.26-5.07) | 0.57 (0.07-4.32) | 2.62 (0.29-23.71) |
| Unadjusted | 1 - pre closure | Men | 41 - 50 | Hospitalisation | 1 | 6.02 (2.89-12.57) | 2.25 (1.02-5.00) | 0.53 (0.12-2.36) | - |
| Unadjusted | 1 - pre closure | Men | 51 - 65 | Hospitalisation | 1 | 1.42 (0.75-2.69) | 1.85 (1.21-2.84) | 0.62 (0.27-1.43) | 0.74 (0.17-3.17) |
| Unadjusted | 1 - pre closure | Women | 21 - 30 | Hospitalisation | 1 | 2.98 (0.92-9.67) | 0.84 (0.10-6.69) | - | 2.13 (0.44-10.33) |
| Unadjusted | 1 - pre closure | Women | 31 - 40 | Hospitalisation | 1 | 2.52 (0.99-6.41) | - | - | 1.23 (0.27-5.50) |
| Unadjusted | 1 - pre closure | Women | 41 - 50 | Hospitalisation | 1 | 3.32 (1.90-5.79) | 1.97 (0.42-9.24) | 2.16 (0.45-10.30) | 0.92 (0.21-3.94) |
| Unadjusted | 1 - pre closure | Women | 51 - 65 | Hospitalisation | 1 | 1.33 (0.80-2.19) | 0.83 (0.19-3.57) | 0.84 (0.26-2.75) | 0.51 (0.16-1.67) |
| Unadjusted | 2 - closed (before phase 3 easing) | Men | 21 - 30 | Hospitalisation | 1 | 6.93 (1.21-39.89) | 9.32 (3.13-27.72) | 1.19 (0.14-10.14) | - |
| Unadjusted | 2 - closed (before phase 3 easing) | Men | 31 - 40 | Hospitalisation | 1 | 5.58 (1.82-17.14) | 9.14 (3.42-24.44) | - | - |
| Unadjusted | 2 - closed (before phase 3 easing) | Men | 41 - 50 | Hospitalisation | 1 | 6.07 (3.20-11.53) | 1.91 (0.98-3.74) | 0.52 (0.12-2.24) | - |
| Unadjusted | 2 - closed (before phase 3 easing) | Men | 51 - 65 | Hospitalisation | 1 | 3.36 (2.14-5.28) | 2.09 (1.44-3.04) | 0.87 (0.44-1.74) | 0.35 (0.05-2.60) |
| Unadjusted | 2 - closed (before phase 3 easing) | Women | 21 - 30 | Hospitalisation | 1 | 6.56 (3.15-13.66) | 1.44 (0.31-6.76) | - | - |
| Unadjusted | 2 - closed (before phase 3 easing) | Women | 31 - 40 | Hospitalisation | 1 | 3.50 (1.91-6.41) | 0.81 (0.11-6.25) | - | 0.56 (0.13-2.34) |
| Unadjusted | 2 - closed (before phase 3 easing) | Women | 41 - 50 | Hospitalisation | 1 | 3.62 (2.42-5.43) | 3.01 (1.00-9.06) | - | 0.32 (0.08-1.31) |

| Adjustment | Time period | Sex | Age group (years) | Outcome | Neither | Healthcare worker | Household member of healthcare worker | Household member of teacher | Teacher |
|---|---|---|---|---|---|---|---|---|---|
| Unadjusted | 2 - closed (before phase 3 easing) | Women | 51 - 65 | Hospitalisation | 1 | 2.66 (1.97-3.58) | 1.74 (0.85-3.56) | 0.90 (0.38-2.12) | - |
| Unadjusted | 2 - closed (phase 3 easing) | Men | 21 - 30 | Hospitalisation | 1 | - | - | - | - |
| Unadjusted | 2 - closed (phase 3 easing) | Men | 31 - 40 | Hospitalisation | 1 | - | - | - | - |
| Unadjusted | 2 - closed (phase 3 easing) | Men | 41 - 50 | Hospitalisation | 1 | - | - | - | - |
| Unadjusted | 2 - closed (phase 3 easing) | Men | 51 - 65 | Hospitalisation | 1 | - | - | - | - |
| Unadjusted | 2 - closed (phase 3 easing) | Women | 21 - 30 | Hospitalisation | 1 | - | - | - | - |
| Unadjusted | 2 - closed (phase 3 easing) | Women | 31 - 40 | Hospitalisation | 1 | - | - | - | - |
| Unadjusted | 2 - closed (phase 3 easing) | Women | 41 - 50 | Hospitalisation | 1 | - | - | - | - |
| Unadjusted | 2 - closed (phase 3 easing) | Women | 51 - 65 | Hospitalisation | 1 | - | - | - | - |
| Unadjusted | 3 - reopened | Men | 21 - 30 | Hospitalisation | 1 | 14.41 (4.17-49.82) | 0.44 (0.10-1.86) | 0.82 (0.19-3.49) | 2.47 (0.28-21.97) |
| Unadjusted | 3 - reopened | Men | 31 - 40 | Hospitalisation | 1 | 2.24 (0.73-6.82) | 1.04 (0.41-2.62) | 0.58 (0.18-1.89) | 1.02 (0.24-4.39) |
| Unadjusted | 3 - reopened | Men | 41 - 50 | Hospitalisation | 1 | 1.41 (0.69-2.85) | 1.30 (0.75-2.25) | 1.24 (0.66-2.34) | 1.99 (0.87-4.53) |
| Unadjusted | 3 - reopened | Men | 51 - 65 | Hospitalisation | 1 | 1.66 (1.15-2.40) | 1.19 (0.88-1.62) | 1.11 (0.75-1.65) | 0.26 (0.06-1.04) |
| Unadjusted | 3 - reopened | Women | 21 - 30 | Hospitalisation | 1 | 1.79 (0.97-3.30) | 0.87 (0.30-2.48) | 1.05 (0.31-3.48) | 0.91 (0.39-2.15) |
| Unadjusted | 3 - reopened | Women | 31 - 40 | Hospitalisation | 1 | 1.95 (1.27-3.00) | 1.49 (0.52-4.31) | 0.34 (0.05-2.50) | 1.33 (0.78-2.28) |
| Unadjusted | 3 - reopened | Women | 41 - 50 | Hospitalisation | 1 | 1.89 (1.31-2.74) | 1.13 (0.45-2.86) | 1.28 (0.49-3.29) | 0.95 (0.50-1.77) |

| Adjustment | Time period | Sex | Age group (years) | Outcome | Neither | Healthcare worker | Household member of healthcare worker | Household member of teacher | Teacher |
|---|---|---|---|---|---|---|---|---|---|
| Unadjusted | 3 - reopened | Women | 51 - 65 | Hospitalisation | 1 | 1.49 (1.15-1.92) | 1.87 (1.20-2.93) | 0.90 (0.49-1.68) | 0.57 (0.30-1.09) |
| Adjusted | 1 - pre closure | Men | 21 - 30 | Hospitalisation | 1 | - | - | - | - |
| Adjusted | 1 - pre closure | Men | 31 - 40 | Hospitalisation | 1 | - | - | - | - |
| Adjusted | 1 - pre closure | Men | 41 - 50 | Hospitalisation | 1 | 6.86 (3.15-14.94) | 2.12 (0.93-4.85) | 0.51 (0.11-2.38) | - |
| Adjusted | 1 - pre closure | Men | 51 - 65 | Hospitalisation | 1 | 1.48 (0.77-2.83) | 1.66 (1.07-2.59) | 0.59 (0.25-1.37) | 0.65 (0.15-2.82) |
| Adjusted | 1 - pre closure | Women | 21 - 30 | Hospitalisation | 1 | 5.42 (1.36-21.66) | 1.09 (0.11-10.80) | - | 2.59 (0.44-15.39) |
| Adjusted | 1 - pre closure | Women | 31 - 40 | Hospitalisation | 1 | - | - | - | - |
| Adjusted | 1 - pre closure | Women | 41 - 50 | Hospitalisation | 1 | - | - | - | - |
| Adjusted | 1 - pre closure | Women | 51 - 65 | Hospitalisation | 1 | 1.47 (0.88-2.48) | 0.65 (0.14-2.99) | 0.87 (0.26-2.98) | 0.66 (0.20-2.24) |
| Adjusted | 2 - closed (before phase 3 easing) | Men | 21 - 30 | Hospitalisation | 1 | - | - | - | - |
| Adjusted | 2 - closed (before phase 3 easing) | Men | 31 - 40 | Hospitalisation | 1 | - | - | - | - |
| Adjusted | 2 - closed (before phase 3 easing) | Men | 41 - 50 | Hospitalisation | 1 | 6.23 (3.16-12.28) | 1.58 (0.76-3.29) | 0.68 (0.15-3.06) | - |
| Adjusted | 2 - closed (before phase 3 easing) | Men | 51 - 65 | Hospitalisation | 1 | 3.65 (2.28-5.84) | 2.07 (1.40-3.07) | 0.78 (0.38-1.62) | 0.43 (0.06-3.23) |
| Adjusted | 2 - closed (before phase 3 easing) | Women | 21 - 30 | Hospitalisation | 1 | - | - | - | - |
| Adjusted | 2 - closed (before phase 3 easing) | Women | 31 - 40 | Hospitalisation | 1 | 4.21 (2.20-8.08) | 0.96 (0.12-7.69) | - | 0.63 (0.15-2.72) |
| Adjusted | 2 - closed (before phase 3 easing) | Women | 41 - 50 | Hospitalisation | 1 | 4.81 (3.10-7.45) | 2.10 (0.60-7.39) | - | 0.41 (0.10-1.75) |
| Adjusted | 2 - closed (before phase 3 easing) | Women | 51 - 65 | Hospitalisation | 1 | 3.00 (2.20-4.09) | 1.52 (0.71-3.25) | 0.90 (0.37-2.17) | - |
| Adjusted | 2 - closed (phase 3 easing) | Men | 21 - 30 | Hospitalisation | 1 | - | - | - | - |

| Adjustment | Time period | Sex | Age group (years) | Outcome | Neither | Healthcare worker | Household member of healthcare worker | Household member of teacher | Teacher |
|---|---|---|---|---|---|---|---|---|---|
| Adjusted | 2 - closed (phase 3 easing) | Men | 31 - 40 | Hospitalisation | 1 | - | - | - | - |
| Adjusted | 2 - closed (phase 3 easing) | Men | 51 - 65 | Hospitalisation | 1 | - | - | - | - |
| Adjusted | 2 - closed (phase 3 easing) | Women | 21 - 30 | Hospitalisation | 1 | - | - | - | - |
| Adjusted | 2 - closed (phase 3 easing) | Women | 31 - 40 | Hospitalisation | 1 | - | - | - | - |
| Adjusted | 2 - closed (phase 3 easing) | Women | 41 - 50 | Hospitalisation | 1 | - | - | - | - |
| Adjusted | 3 - reopened | Men | 21 - 30 | Hospitalisation | 1 | 15.32 (3.93-59.82) | 0.40 (0.09-1.82) | 0.77 (0.14-4.35) | 1.31 (0.13-13.39) |
| Adjusted | 3 - reopened | Men | 31 - 40 | Hospitalisation | 1 | 3.05 (0.92-10.04) | 1.04 (0.40-2.70) | 0.64 (0.19-2.10) | 0.99 (0.22-4.49) |
| Adjusted | 3 - reopened | Men | 41 - 50 | Hospitalisation | 1 | 1.67 (0.80-3.49) | 1.06 (0.57-1.95) | 1.39 (0.72-2.71) | 2.51 (1.03-6.12) |
| Adjusted | 3 - reopened | Men | 51 - 65 | Hospitalisation | 1 | 1.64 (1.11-2.43) | 1.00 (0.73-1.38) | 1.01 (0.67-1.54) | 0.18 (0.04-0.77) |
| Adjusted | 3 - reopened | Women | 21 - 30 | Hospitalisation | 1 | 1.76 (0.93-3.32) | 0.87 (0.30-2.57) | 1.29 (0.37-4.48) | 1.00 (0.41-2.45) |
| Adjusted | 3 - reopened | Women | 31 - 40 | Hospitalisation | 1 | 2.16 (1.36-3.41) | 0.90 (0.29-2.83) | 0.29 (0.03-2.61) | 1.42 (0.80-2.51) |
| Adjusted | 3 - reopened | Women | 41 - 50 | Hospitalisation | 1 | 2.13 (1.43-3.17) | 1.12 (0.42-3.00) | 1.51 (0.56-4.10) | 1.04 (0.53-2.02) |
| Adjusted | 3 - reopened | Women | 51 - 65 | Hospitalisation | 1 | 1.62 (1.24-2.12) | 1.39 (0.85-2.26) | 0.68 (0.35-1.30) | 0.72 (0.38-1.40) |
| Unadjusted | 1 - pre closure | Men | 21 - 30 | Any case | 1 | 60.07 (24.47-147.41) | 6.06 (2.98-12.31) | 3.54 (1.09-11.51) | 2.85 (0.52-15.50) |
| Unadjusted | 1 - pre closure | Men | 31 - 40 | Any case | 1 | 26.67 (13.96-50.95) | 7.50 (4.47-12.58) | 0.79 (0.27-2.26) | 1.17 (0.15-9.24) |
| Unadjusted | 1 - pre closure | Men | 41 - 50 | Any case | 1 | 22.54 (14.42-35.24) | 5.54 (3.48-8.83) | 0.40 (0.11-1.46) | 0.58 (0.07-4.54) |

| Adjustment | Time period | Sex | Age group (years) | Outcome | Neither | Healthcare worker | Household member of healthcare worker | Household member of teacher | Teacher |
|---|---|---|---|---|---|---|---|---|---|
| Unadjusted | 1 - pre closure | Men | 51 - 65 | Any case | 1 | 7.21 (5.31-9.78) | 3.20 (2.37-4.31) | 0.61 (0.31-1.18) | 0.52 (0.12-2.19) |
| Unadjusted | 1 - pre closure | Women | 21 - 30 | Any case | 1 | 20.69 (12.99-32.97) | 4.54 (2.15-9.58) | 0.22 (0.02-2.09) | 0.72 (0.17-3.11) |
| Unadjusted | 1 - pre closure | Women | 31 - 40 | Any case | 1 | 12.88 (9.08-18.25) | 3.92 (1.24-12.43) | 1.21 (0.42-3.48) | 0.51 (0.16-1.64) |
| Unadjusted | 1 - pre closure | Women | 41 - 50 | Any case | 1 | 11.41 (8.54-15.24) | 4.45 (2.08-9.54) | 1.73 (0.62-4.82) | 0.44 (0.14-1.40) |
| Unadjusted | 1 - pre closure | Women | 51 - 65 | Any case | 1 | 6.96 (5.58-8.68) | 3.08 (1.69-5.60) | 0.79 (0.33-1.89) | 0.50 (0.20-1.25) |
| Unadjusted | 2 - closed (before phase 3 easing) | Men | 21 - 30 | Any case | 1 | 24.55 (16.45-36.64) | 2.84 (1.92-4.21) | 0.78 (0.36-1.66) | 0.50 (0.07-3.79) |
| Unadjusted | 2 - closed (before phase 3 easing) | Men | 31 - 40 | Any case | 1 | 24.22 (16.83-34.85) | 3.74 (2.60-5.38) | 0.55 (0.25-1.24) | 0.44 (0.11-1.85) |
| Unadjusted | 2 - closed (before phase 3 easing) | Men | 41 - 50 | Any case | 1 | 22.04 (16.35-29.72) | 3.03 (2.17-4.24) | 0.52 (0.24-1.12) | 1.21 (0.42-3.47) |
| Unadjusted | 2 - closed (before phase 3 easing) | Men | 51 - 65 | Any case | 1 | 10.47 (8.37-13.10) | 2.90 (2.32-3.63) | 1.14 (0.77-1.71) | 0.68 (0.25-1.88) |
| Unadjusted | 2 - closed (before phase 3 easing) | Women | 21 - 30 | Any case | 1 | 13.31 (11.35-15.62) | 1.49 (1.03-2.15) | 0.60 (0.32-1.10) | 0.27 (0.12-0.60) |
| Unadjusted | 2 - closed (before phase 3 easing) | Women | 31 - 40 | Any case | 1 | 9.17 (7.87-10.68) | 1.24 (0.68-2.25) | 0.86 (0.43-1.72) | 0.36 (0.19-0.65) |
| Unadjusted | 2 - closed (before phase 3 easing) | Women | 41 - 50 | Any case | 1 | 7.46 (6.51-8.54) | 2.92 (1.95-4.39) | 0.68 (0.34-1.36) | 0.17 (0.07-0.37) |
| Unadjusted | 2 - closed (before phase 3 easing) | Women | 51 - 65 | Any case | 1 | 6.63 (5.91-7.44) | 2.50 (1.85-3.38) | 0.64 (0.41-1.01) | 0.09 (0.03-0.27) |
| Unadjusted | 2 - closed (phase 3 easing) | Men | 21 - 30 | Any case | 1 | 0.58 (0.14-2.40) | 1.02 (0.59-1.76) | 1.07 (0.55-2.08) | 1.36 (0.47-3.90) |
| Unadjusted | 2 - closed (phase 3 easing) | Men | 31 - 40 | Any case | 1 | 1.27 (0.37-4.33) | 0.79 (0.31-1.99) | 1.07 (0.42-2.73) | - |
| Unadjusted | 2 - closed (phase 3 easing) | Men | 41 - 50 | Any case | 1 | 2.01 (0.82-4.92) | 1.06 (0.37-3.02) | 1.26 (0.48-3.27) | 3.34 (1.03-10.85) |

| Adjustment | Time period | Sex | Age group (years) | Outcome | Neither | Healthcare worker | Household member of healthcare worker | Household member of teacher | Teacher |
|---|---|---|---|---|---|---|---|---|---|
| Unadjusted | 2 - closed (phase 3 easing) | Men | 51 - 65 | Any case | 1 | 0.36 (0.05-2.72) | 1.58 (0.77-3.26) | 1.74 (0.72-4.18) | 3.24 (0.65-16.13) |
| Unadjusted | 2 - closed (phase 3 easing) | Women | 21 - 30 | Any case | 1 | 1.75 (1.02-3.00) | 1.13 (0.54-2.37) | 1.95 (0.94-4.06) | 1.16 (0.49-2.75) |
| Unadjusted | 2 - closed (phase 3 easing) | Women | 31 - 40 | Any case | 1 | 0.52 (0.21-1.33) | 1.15 (0.14-9.21) | - | 1.56 (0.76-3.22) |
| Unadjusted | 2 - closed (phase 3 easing) | Women | 41 - 50 | Any case | 1 | 1.02 (0.46-2.25) | - | 1.00 (0.12-8.11) | 0.98 (0.38-2.53) |
| Unadjusted | 2 - closed (phase 3 easing) | Women | 51 - 65 | Any case | 1 | 1.13 (0.64-2.00) | 0.32 (0.04-2.38) | 1.38 (0.47-4.00) | 1.77 (0.74-4.24) |
| Unadjusted | 3 - reopened | Men | 21 - 30 | Any case | 1 | 3.33 (2.80-3.96) | 1.37 (1.22-1.53) | 1.27 (1.11-1.46) | 1.97 (1.56-2.49) |
| Unadjusted | 3 - reopened | Men | 31 - 40 | Any case | 1 | 2.78 (2.38-3.26) | 1.49 (1.32-1.69) | 1.27 (1.11-1.46) | 1.94 (1.60-2.35) |
| Unadjusted | 3 - reopened | Men | 41 - 50 | Any case | 1 | 2.47 (2.13-2.86) | 1.74 (1.54-1.96) | 1.34 (1.16-1.55) | 1.89 (1.53-2.33) |
| Unadjusted | 3 - reopened | Men | 51 - 65 | Any case | 1 | 2.60 (2.32-2.92) | 1.73 (1.57-1.90) | 1.37 (1.21-1.54) | 1.39 (1.10-1.76) |
| Unadjusted | 3 - reopened | Women | 21 - 30 | Any case | 1 | 2.63 (2.43-2.85) | 1.51 (1.34-1.70) | 1.06 (0.91-1.23) | 1.47 (1.31-1.64) |
| Unadjusted | 3 - reopened | Women | 31 - 40 | Any case | 1 | 2.38 (2.21-2.56) | 1.21 (0.97-1.51) | 0.83 (0.65-1.06) | 1.31 (1.18-1.45) |
| Unadjusted | 3 - reopened | Women | 41 - 50 | Any case | 1 | 2.16 (2.00-2.33) | 1.28 (1.04-1.58) | 1.10 (0.87-1.38) | 1.30 (1.16-1.46) |
| Unadjusted | 3 - reopened | Women | 51 - 65 | Any case | 1 | 2.25 (2.11-2.41) | 1.65 (1.43-1.92) | 1.27 (1.10-1.48) | 1.21 (1.06-1.38) |
| Adjusted | 1 - pre closure | Men | 21 - 30 | Any case | 1 | 71.04 (27.27-185.04) | 6.18 (2.90-13.18) | 3.51 (1.07-11.53) | 4.08 (0.65-25.65) |
| Adjusted | 1 - pre closure | Men | 31 - 40 | Any case | 1 | 27.11 (13.79-53.28) | 6.76 (3.96-11.54) | 0.72 (0.26-2.02) | 1.03 (0.13-8.31) |
| Adjusted | 1 - pre closure | Men | 41 - 50 | Any case | 1 | 23.94 (15.07-38.04) | 5.31 (3.29-8.58) | 0.35 (0.09-1.30) | 0.55 (0.07-4.29) |

| Adjustment | Time period | Sex | Age group (years) | Outcome | Neither | Healthcare worker | Household member of healthcare worker | Household member of teacher | Teacher |
|---|---|---|---|---|---|---|---|---|---|
| Adjusted | 1 - pre closure | Men | 51 - 65 | Any case | 1 | 7.29 (5.32-9.98) | 2.92 (2.15-3.98) | 0.56 (0.29-1.09) | 0.48 (0.11-2.05) |
| Adjusted | 1 - pre closure | Women | 21 - 30 | Any case | 1 | 23.69 (14.42-38.91) | 4.90 (2.29-10.51) | 0.32 (0.03-3.28) | 0.72 (0.16-3.19) |
| Adjusted | 1 - pre closure | Women | 31 - 40 | Any case | 1 | 13.10 (9.11-18.83) | 3.44 (1.04-11.37) | 1.28 (0.44-3.73) | 0.51 (0.16-1.64) |
| Adjusted | 1 - pre closure | Women | 41 - 50 | Any case | 1 | 11.42 (8.51-15.32) | 4.15 (1.90-9.07) | 1.72 (0.62-4.78) | 0.45 (0.14-1.47) |
| Adjusted | 1 - pre closure | Women | 51 - 65 | Any case | 1 | 7.41 (5.89-9.33) | 3.04 (1.63-5.67) | 0.70 (0.28-1.72) | 0.57 (0.23-1.42) |
| Adjusted | 2 - closed (before phase 3 easing) | Men | 21 - 30 | Any case | 1 | 25.00 (16.65-37.55) | 2.47 (1.66-3.69) | 0.69 (0.32-1.51) | 0.49 (0.06-3.73) |
| Adjusted | 2 - closed (before phase 3 easing) | Men | 31 - 40 | Any case | 1 | 25.72 (17.65-37.49) | 3.37 (2.32-4.91) | 0.55 (0.24-1.23) | 0.47 (0.11-1.97) |
| Adjusted | 2 - closed (before phase 3 easing) | Men | 41 - 50 | Any case | 1 | 23.01 (16.91-31.30) | 2.55 (1.81-3.61) | 0.50 (0.23-1.08) | 1.22 (0.41-3.62) |
| Adjusted | 2 - closed (before phase 3 easing) | Men | 51 - 65 | Any case | 1 | 10.66 (8.46-13.44) | 2.60 (2.06-3.28) | 1.04 (0.69-1.56) | 0.75 (0.27-2.10) |
| Adjusted | 2 - closed (before phase 3 easing) | Women | 21 - 30 | Any case | 1 | 13.58 (11.54-15.97) | 1.40 (0.96-2.02) | 0.57 (0.31-1.06) | 0.26 (0.12-0.60) |
| Adjusted | 2 - closed (before phase 3 easing) | Women | 31 - 40 | Any case | 1 | 9.74 (8.32-11.39) | 1.24 (0.68-2.27) | 0.88 (0.43-1.80) | 0.40 (0.22-0.73) |
| Adjusted | 2 - closed (before phase 3 easing) | Women | 41 - 50 | Any case | 1 | 7.70 (6.70-8.85) | 2.89 (1.92-4.37) | 0.65 (0.32-1.33) | 0.18 (0.08-0.41) |
| Adjusted | 2 - closed (before phase 3 easing) | Women | 51 - 65 | Any case | 1 | 6.76 (6.01-7.60) | 2.39 (1.76-3.24) | 0.63 (0.40-0.99) | 0.10 (0.03-0.30) |
| Adjusted | 2 - closed (phase 3 easing) | Men | 21 - 30 | Any case | 1 | 0.53 (0.13-2.26) | 0.94 (0.54-1.63) | 0.92 (0.47-1.81) | 1.27 (0.44-3.71) |
| Adjusted | 2 - closed (phase 3 easing) | Men | 31 - 40 | Any case | 1 | 1.29 (0.37-4.45) | 0.72 (0.28-1.83) | 0.98 (0.38-2.54) | - |
| Adjusted | 2 - closed (phase 3 easing) | Men | 51 - 65 | Any case | 1 | 0.36 (0.05-2.78) | 1.20 (0.57-2.53) | 1.27 (0.51-3.17) | 2.42 (0.45-12.93) |

| **Adjustment** | **Time period** | **Sex** | **Age group (years)** | **Outcome** | **Neither** | **Healthcare worker** | **Household member of healthcare worker** | **Household member of teacher** | **Teacher** |
|---|---|---|---|---|---|---|---|---|---|
| Adjusted | 2 - closed (phase 3 easing) | Women | 21 - 30 | Any case | 1 | 1.76 (1.02-3.06) | 1.00 (0.47-2.11) | 1.67 (0.79-3.53) | 1.16 (0.49-2.77) |
| Adjusted | 2 - closed (phase 3 easing) | Women | 31 - 40 | Any case | 1 | 0.50 (0.19-1.31) | 0.86 (0.10-7.34) | - | 1.61 (0.75-3.44) |
| Adjusted | 2 - closed (phase 3 easing) | Women | 41 - 50 | Any case | 1 | 0.93 (0.41-2.07) | - | 1.34 (0.16-11.44) | 0.82 (0.31-2.17) |
| Adjusted | 3 - reopened | Men | 21 - 30 | Any case | 1 | 3.38 (2.83-4.02) | 1.19 (1.06-1.34) | 1.14 (0.99-1.31) | 1.96 (1.54-2.49) |
| Adjusted | 3 - reopened | Men | 31 - 40 | Any case | 1 | 2.74 (2.33-3.23) | 1.33 (1.18-1.51) | 1.16 (1.01-1.33) | 1.94 (1.59-2.35) |
| Adjusted | 3 - reopened | Men | 41 - 50 | Any case | 1 | 2.35 (2.02-2.75) | 1.35 (1.19-1.53) | 1.07 (0.92-1.25) | 1.95 (1.56-2.43) |
| Adjusted | 3 - reopened | Men | 51 - 65 | Any case | 1 | 2.50 (2.22-2.83) | 1.30 (1.17-1.43) | 0.99 (0.87-1.13) | 1.25 (0.98-1.59) |
| Adjusted | 3 - reopened | Women | 21 - 30 | Any case | 1 | 2.69 (2.48-2.92) | 1.29 (1.14-1.46) | 0.92 (0.79-1.07) | 1.51 (1.35-1.69) |
| Adjusted | 3 - reopened | Women | 31 - 40 | Any case | 1 | 2.42 (2.24-2.62) | 1.04 (0.83-1.30) | 0.74 (0.57-0.95) | 1.41 (1.27-1.57) |
| Adjusted | 3 - reopened | Women | 41 - 50 | Any case | 1 | 2.14 (1.98-2.32) | 1.01 (0.81-1.26) | 0.87 (0.69-1.10) | 1.26 (1.12-1.43) |
| Adjusted | 3 - reopened | Women | 51 - 65 | Any case | 1 | 2.24 (2.09-2.40) | 1.22 (1.05-1.42) | 0.91 (0.78-1.06) | 1.16 (1.01-1.33) |

Table S4b Rate ratios for any case and hospitalisation with COVID-19 for teachers by sector, stratified by time period, age and sex

| Adjustment | Time period | Sex | Age group (years) | Outcome | Neither | Nursery/Primary or Nursery | Primary | Secondary | Teacher in other sector |
|---|---|---|---|---|---|---|---|---|---|
| Unadjusted | 1 - pre closure | Men | 21 - 30 | Any case | 1 | - | 15.94 (0.91-278.20) | - | 11.01 (0.18-682.10) |
| Adjusted | 1 - pre closure | Men | 21 - 30 | Any case | 1 | - | 28.54 (1.49-545.24) | - | 14.72 (0.10-2081.40) |
| Unadjusted | 1 - pre closure | Men | 21 - 30 | Hospitalisation | 1 | - | - | - | - |
| Adjusted | 1 - pre closure | Men | 21 - 30 | Hospitalisation | 1 | - | - | - | - |
| Unadjusted | 1 - pre closure | Men | 31 - 40 | Any case | 1 | - | - | 1.81 (0.22-14.98) | - |
| Adjusted | 1 - pre closure | Men | 31 - 40 | Any case | 1 | - | - | 1.53 (0.18-13.21) | - |
| Unadjusted | 1 - pre closure | Men | 31 - 40 | Hospitalisation | 1 | - | - | 3.18 (0.33-30.75) | - |
| Adjusted | 1 - pre closure | Men | 31 - 40 | Hospitalisation | 1 | - | - | - | - |
| Unadjusted | 1 - pre closure | Men | 41 - 50 | Any case | 1 | - | - | - | 1.55 (0.15-16.04) |
| Adjusted | 1 - pre closure | Men | 41 - 50 | Any case | 1 | - | - | - | 2.02 (0.20-20.01) |
| Unadjusted | 1 - pre closure | Men | 41 - 50 | Hospitalisation | 1 | - | - | - | - |
| Adjusted | 1 - pre closure | Men | 41 - 50 | Hospitalisation | 1 | - | - | - | - |
| Unadjusted | 1 - pre closure | Men | 51 - 65 | Any case | 1 | - | - | 0.50 (0.07-3.69) | 0.92 (0.10-8.32) |
| Adjusted | 1 - pre closure | Men | 51 - 65 | Any case | 1 | - | - | 0.46 (0.06-3.43) | 0.89 (0.10-8.40) |
| Unadjusted | 1 - pre closure | Men | 51 - 65 | Hospitalisation | 1 | - | - | 0.82 (0.11-6.38) | 1.26 (0.14-11.17) |
| Adjusted | 1 - pre closure | Men | 51 - 65 | Hospitalisation | 1 | - | - | 0.70 (0.09-5.55) | 1.22 (0.13-11.26) |
| Unadjusted | 1 - pre closure | Women | 21 - 30 | Any case | 1 | - | 2.48 (0.54-11.48) | - | - |
| Adjusted | 1 - pre closure | Women | 21 - 30 | Any case | 1 | - | 2.25 (0.47-10.73) | - | - |

| Adjustment | Time period | Sex | Age group (years) | Outcome | Neither | Nursery/Primary or Nursery | Primary | Secondary | Teacher in other sector |
|---|---|---|---|---|---|---|---|---|---|
| Unadjusted | 1 - pre closure | Women | 21 - 30 | Hospitalisation | 1 | - | 4.85 (0.88-26.64) | - | - |
| Adjusted | 1 - pre closure | Women | 21 - 30 | Hospitalisation | 1 | - | 5.13 (0.72-36.36) | - | - |
| Unadjusted | 1 - pre closure | Women | 31 - 40 | Any case | 1 | 0.51 (0.07-3.87) | - | 0.46 (0.06-3.39) | 1.78 (0.23-13.97) |
| Adjusted | 1 - pre closure | Women | 31 - 40 | Any case | 1 | 0.49 (0.06-3.69) | - | 0.49 (0.07-3.68) | 2.01 (0.25-15.92) |
| Unadjusted | 1 - pre closure | Women | 31 - 40 | Hospitalisation | 1 | - | - | 1.82 (0.22-15.24) | 3.79 (0.39-36.90) |
| Adjusted | 1 - pre closure | Women | 31 - 40 | Hospitalisation | 1 | - | - | - | - |
| Unadjusted | 1 - pre closure | Women | 41 - 50 | Any case | 1 | - | 0.90 (0.12-6.75) | 1.00 (0.23-4.33) | - |
| Adjusted | 1 - pre closure | Women | 41 - 50 | Any case | 1 | - | 0.88 (0.12-6.68) | 0.99 (0.23-4.37) | - |
| Unadjusted | 1 - pre closure | Women | 41 - 50 | Hospitalisation | 1 | - | 2.45 (0.28-21.13) | 1.61 (0.20-13.14) | - |
| Adjusted | 1 - pre closure | Women | 41 - 50 | Hospitalisation | 1 | - | - | - | - |
| Unadjusted | 1 - pre closure | Women | 51 - 65 | Any case | 1 | - | - | 1.09 (0.39-3.09) | 0.41 (0.05-3.14) |
| Adjusted | 1 - pre closure | Women | 51 - 65 | Any case | 1 | - | - | 1.18 (0.41-3.36) | 0.41 (0.05-3.15) |
| Unadjusted | 1 - pre closure | Women | 51 - 65 | Hospitalisation | 1 | - | - | 0.89 (0.21-3.80) | 0.74 (0.09-6.05) |
| Adjusted | 1 - pre closure | Women | 51 - 65 | Hospitalisation | 1 | - | - | 1.15 (0.26-5.13) | 0.75 (0.09-6.41) |
| Unadjusted | 2 - closed (before phase 3 easing) | Men | 21 - 30 | Any case | 1 | 2.39 (0.28-20.66) | - | - | - |
| Adjusted | 2 - closed (before phase 3 easing) | Men | 21 - 30 | Any case | 1 | 2.10 (0.23-19.30) | - | - | - |
| Unadjusted | 2 - closed (before phase 3 easing) | Men | 21 - 30 | Hospitalisation | 1 | - | - | - | - |
| Adjusted | 2 - closed (before phase 3 easing) | Men | 21 - 30 | Hospitalisation | 1 | - | - | - | - |

| Adjustment | Time period | Sex | Age group (years) | Outcome | Neither | Nursery/Primary or Nursery | Primary | Secondary | Teacher in other sector |
|---|---|---|---|---|---|---|---|---|---|
| Unadjusted | 2 - closed (before phase 3 easing) | Men | 31 - 40 | Any case | 1 | - | - | 0.78 (0.18-3.33) | - |
| Adjusted | 2 - closed (before phase 3 easing) | Men | 31 - 40 | Any case | 1 | - | - | 0.82 (0.19-3.56) | - |
| Unadjusted | 2 - closed (before phase 3 easing) | Men | 31 - 40 | Hospitalisation | 1 | - | - | - | - |
| Adjusted | 2 - closed (before phase 3 easing) | Men | 31 - 40 | Hospitalisation | 1 | - | - | - | - |
| Unadjusted | 2 - closed (before phase 3 easing) | Men | 41 - 50 | Any case | 1 | - | 2.10 (0.22-20.20) | 0.49 (0.06-3.67) | 4.44 (0.87-22.69) |
| Adjusted | 2 - closed (before phase 3 easing) | Men | 41 - 50 | Any case | 1 | - | 2.05 (0.19-21.54) | 0.46 (0.06-3.62) | 5.77 (1.13-29.53) |
| Unadjusted | 2 - closed (before phase 3 easing) | Men | 41 - 50 | Hospitalisation | 1 | - | - | - | - |
| Adjusted | 2 - closed (before phase 3 easing) | Men | 41 - 50 | Hospitalisation | 1 | - | - | - | - |
| Unadjusted | 2 - closed (before phase 3 easing) | Men | 51 - 65 | Any case | 1 | - | 3.47 (0.38-32.02) | 0.81 (0.25-2.63) | - |
| Adjusted | 2 - closed (before phase 3 easing) | Men | 51 - 65 | Any case | 1 | - | 3.78 (0.40-35.40) | 0.97 (0.29-3.18) | - |
| Unadjusted | 2 - closed (before phase 3 easing) | Men | 51 - 65 | Hospitalisation | 1 | - | - | 0.55 (0.07-4.13) | - |
| Adjusted | 2 - closed (before phase 3 easing) | Men | 51 - 65 | Hospitalisation | 1 | - | - | 0.65 (0.08-5.02) | - |
| Unadjusted | 2 - closed (before phase 3 easing) | Women | 21 - 30 | Any case | 1 | 0.17 (0.02-1.24) | 0.35 (0.11-1.12) | 0.14 (0.02-1.01) | 0.88 (0.12-6.72) |
| Adjusted | 2 - closed (before phase 3 easing) | Women | 21 - 30 | Any case | 1 | 0.17 (0.02-1.23) | 0.36 (0.11-1.14) | 0.14 (0.02-0.99) | 0.85 (0.11-6.52) |
| Unadjusted | 2 - closed (before phase 3 easing) | Women | 21 - 30 | Hospitalisation | 1 | - | - | - | - |
| Adjusted | 2 - closed (before phase 3 easing) | Women | 21 - 30 | Hospitalisation | 1 | - | - | - | - |
| Unadjusted | 2 - closed (before phase 3 easing) | Women | 31 - 40 | Any case | 1 | 0.40 (0.13-1.25) | 0.42 (0.15-1.13) | 0.29 (0.09-0.92) | 0.29 (0.04-2.10) |

| Adjustment | Time period | Sex | Age group (years) | Outcome | Neither | Nursery/Primary or Nursery | Primary | Secondary | Teacher in other sector |
|---|---|---|---|---|---|---|---|---|---|
| Adjusted | 2 - closed (before phase 3 easing) | Women | 31 - 40 | Any case | 1 | 0.44 (0.14-1.39) | 0.47 (0.17-1.29) | 0.32 (0.10-1.01) | 0.33 (0.05-2.41) |
| Unadjusted | 2 - closed (before phase 3 easing) | Women | 31 - 40 | Hospitalisation | 1 | 1.44 (0.17-11.92) | - | 0.88 (0.11-6.75) | - |
| Adjusted | 2 - closed (before phase 3 easing) | Women | 31 - 40 | Hospitalisation | 1 | 2.43 (0.28-21.21) | - | 0.85 (0.10-7.04) | - |
| Unadjusted | 2 - closed (before phase 3 easing) | Women | 41 - 50 | Any case | 1 | 0.28 (0.09-0.90) | 0.12 (0.02-0.84) | - | 0.35 (0.09-1.45) |
| Adjusted | 2 - closed (before phase 3 easing) | Women | 41 - 50 | Any case | 1 | 0.33 (0.10-1.03) | 0.12 (0.02-0.89) | - | 0.39 (0.10-1.61) |
| Unadjusted | 2 - closed (before phase 3 easing) | Women | 41 - 50 | Hospitalisation | 1 | 0.97 (0.12-7.82) | - | - | 1.00 (0.12-8.03) |
| Adjusted | 2 - closed (before phase 3 easing) | Women | 41 - 50 | Hospitalisation | 1 | 1.08 (0.11-10.17) | - | - | 1.20 (0.14-9.90) |
| Unadjusted | 2 - closed (before phase 3 easing) | Women | 51 - 65 | Any case | 1 | 0.10 (0.01-0.75) | - | 0.09 (0.01-0.63) | 0.16 (0.02-1.15) |
| Adjusted | 2 - closed (before phase 3 easing) | Women | 51 - 65 | Any case | 1 | 0.12 (0.02-0.87) | - | 0.10 (0.01-0.71) | 0.17 (0.02-1.21) |
| Unadjusted | 2 - closed (before phase 3 easing) | Women | 51 - 65 | Hospitalisation | 1 | - | - | - | - |
| Adjusted | 2 - closed (before phase 3 easing) | Women | 51 - 65 | Hospitalisation | 1 | - | - | - | - |
| Unadjusted | 2 - closed (phase 3 easing) | Men | 21 - 30 | Any case | 1 | - | 6.52 (1.09-39.13) | 0.51 (0.07-3.84) | 4.89 (0.44-54.17) |
| Adjusted | 2 - closed (phase 3 easing) | Men | 21 - 30 | Any case | 1 | - | 4.75 (0.73-31.07) | 0.51 (0.07-3.87) | 5.24 (0.46-59.86) |
| Unadjusted | 2 - closed (phase 3 easing) | Men | 21 - 30 | Hospitalisation | 1 | - | - | - | - |
| Adjusted | 2 - closed (phase 3 easing) | Men | 21 - 30 | Hospitalisation | 1 | - | - | - | - |
| Unadjusted | 2 - closed (phase 3 easing) | Men | 31 - 40 | Any case | 1 | - | - | - | - |
| Adjusted | 2 - closed (phase 3 easing) | Men | 31 - 40 | Any case | 1 | - | - | - | - |

| Adjustment | Time period | Sex | Age group (years) | Outcome | Neither | Nursery/Primary or Nursery | Primary | Secondary | Teacher in other sector |
|---|---|---|---|---|---|---|---|---|---|
| Unadjusted | 2 - closed (phase 3 easing) | Men | 31 - 40 | Hospitalisation | 1 | - | - | - | - |
| Adjusted | 2 - closed (phase 3 easing) | Men | 31 - 40 | Hospitalisation | 1 | - | - | - | - |
| Unadjusted | 2 - closed (phase 3 easing) | Men | 41 - 50 | Any case | 1 | - | 5.37 (0.48-59.70) | 3.51 (0.70-17.53) | 2.78 (0.28-27.87) |
| Unadjusted | 2 - closed (phase 3 easing) | Men | 41 - 50 | Hospitalisation | 1 | - | - | - | - |
| Unadjusted | 2 - closed (phase 3 easing) | Men | 51 - 65 | Any case | 1 | NA | NA | - | 20.83 (1.89-230.07) |
| Adjusted | 2 - closed (phase 3 easing) | Men | 51 - 65 | Any case | 1 | NA | NA | - | 13.09 (1.01-169.21) |
| Unadjusted | 2 - closed (phase 3 easing) | Men | 51 - 65 | Hospitalisation | 1 | NA | NA | - | - |
| Adjusted | 2 - closed (phase 3 easing) | Men | 51 - 65 | Hospitalisation | 1 | NA | NA | - | - |
| Unadjusted | 2 - closed (phase 3 easing) | Women | 21 - 30 | Any case | 1 | 1.44 (0.32-6.42) | 2.89 (0.79-10.66) | 0.46 (0.06-3.44) | - |
| Adjusted | 2 - closed (phase 3 easing) | Women | 21 - 30 | Any case | 1 | 1.42 (0.31-6.44) | 3.13 (0.83-11.74) | 0.45 (0.06-3.37) | - |
| Unadjusted | 2 - closed (phase 3 easing) | Women | 21 - 30 | Hospitalisation | 1 | - | - | - | - |
| Adjusted | 2 - closed (phase 3 easing) | Women | 21 - 30 | Hospitalisation | 1 | - | - | - | - |
| Unadjusted | 2 - closed (phase 3 easing) | Women | 31 - 40 | Any case | 1 | 1.39 (0.41-4.69) | 1.56 (0.35-6.98) | 1.68 (0.48-5.92) | 1.89 (0.22-16.26) |
| Adjusted | 2 - closed (phase 3 easing) | Women | 31 - 40 | Any case | 1 | 1.42 (0.41-4.94) | 1.96 (0.42-9.12) | 1.78 (0.47-6.70) | 1.27 (0.13-12.36) |
| Unadjusted | 2 - closed (phase 3 easing) | Women | 31 - 40 | Hospitalisation | 1 | - | - | - | - |
| Adjusted | 2 - closed (phase 3 easing) | Women | 31 - 40 | Hospitalisation | 1 | - | - | - | - |
| Unadjusted | 2 - closed (phase 3 easing) | Women | 41 - 50 | Any case | 1 | 0.61 (0.08-4.73) | - | 0.87 (0.11-6.72) | 2.64 (0.69-10.10) |

| Adjustment | Time period | Sex | Age group (years) | Outcome | Neither | Nursery/Primary or Nursery | Primary | Secondary | Teacher in other sector |
|---|---|---|---|---|---|---|---|---|---|
| Adjusted | 2 - closed (phase 3 easing) | Women | 41 - 50 | Any case | 1 | 0.48 (0.06-3.85) | - | 0.66 (0.08-5.37) | 2.08 (0.51-8.48) |
| Unadjusted | 2 - closed (phase 3 easing) | Women | 41 - 50 | Hospitalisation | 1 | - | - | - | - |
| Adjusted | 2 - closed (phase 3 easing) | Women | 41 - 50 | Hospitalisation | 1 | - | - | - | - |
| Unadjusted | 2 - closed (phase 3 easing) | Women | 51 - 65 | Any case | 1 | 1.08 (0.13-8.71) | 0.76 (0.10-5.78) | 1.77 (0.21-14.95) | 5.56 (1.31-23.66) |
| Unadjusted | 2 - closed (phase 3 easing) | Women | 51 - 65 | Hospitalisation | 1 | - | - | - | - |
| Unadjusted | 3 - reopened | Men | 21 - 30 | Any case | 1 | 2.39 (1.33-4.30) | 1.70 (0.94-3.07) | 1.89 (1.40-2.56) | 2.56 (1.17-5.58) |
| Adjusted | 3 - reopened | Men | 21 - 30 | Any case | 1 | 2.21 (1.21-4.02) | 1.83 (1.00-3.34) | 1.86 (1.37-2.53) | 2.61 (1.18-5.79) |
| Unadjusted | 3 - reopened | Men | 21 - 30 | Hospitalisation | 1 | - | - | 2.45 (0.28-21.78) | - |
| Adjusted | 3 - reopened | Men | 21 - 30 | Hospitalisation | 1 | - | - | 1.28 (0.12-13.11) | - |
| Unadjusted | 3 - reopened | Men | 31 - 40 | Any case | 1 | 1.70 (1.01-2.85) | 2.06 (1.26-3.38) | 1.95 (1.52-2.50) | 2.04 (1.20-3.44) |
| Adjusted | 3 - reopened | Men | 31 - 40 | Any case | 1 | 1.70 (1.01-2.89) | 2.00 (1.20-3.34) | 1.94 (1.51-2.51) | 2.12 (1.23-3.64) |
| Unadjusted | 3 - reopened | Men | 31 - 40 | Hospitalisation | 1 | 1.97 (0.23-16.91) | - | 0.82 (0.11-6.31) | - |
| Adjusted | 3 - reopened | Men | 31 - 40 | Hospitalisation | 1 | 2.46 (0.27-22.28) | - | 0.72 (0.09-5.99) | - |
| Unadjusted | 3 - reopened | Men | 41 - 50 | Any case | 1 | 1.61 (0.82-3.14) | 3.60 (2.06-6.29) | 1.56 (1.18-2.07) | 2.43 (1.54-3.83) |
| Adjusted | 3 - reopened | Men | 41 - 50 | Any case | 1 | 1.83 (0.92-3.64) | 3.24 (1.80-5.83) | 1.64 (1.22-2.21) | 2.41 (1.49-3.89) |
| Unadjusted | 3 - reopened | Men | 41 - 50 | Hospitalisation | 1 | 2.03 (0.24-17.42) | - | 1.76 (0.60-5.19) | 4.02 (0.78-20.74) |
| Adjusted | 3 - reopened | Men | 41 - 50 | Hospitalisation | 1 | 3.19 (0.34-30.01) | - | 1.94 (0.59-6.35) | 4.79 (0.90-25.41) |

| Adjustment | Time period | Sex | Age group (years) | Outcome | Neither | Nursery/Primary or Nursery | Primary | Secondary | Teacher in other sector |
|---|---|---|---|---|---|---|---|---|---|
| Unadjusted | 3 - reopened | Men | 51 - 65 | Any case | 1 | 2.44 (1.07-5.57) | 0.47 (0.11-1.93) | 1.51 (1.15-1.99) | 1.09 (0.63-1.90) |
| Adjusted | 3 - reopened | Men | 51 - 65 | Any case | 1 | 2.39 (1.01-5.64) | 0.38 (0.09-1.59) | 1.31 (0.98-1.74) | 1.12 (0.63-1.99) |
| Unadjusted | 3 - reopened | Men | 51 - 65 | Hospitalisation | 1 | - | - | 0.39 (0.09-1.60) | - |
| Adjusted | 3 - reopened | Men | 51 - 65 | Hospitalisation | 1 | - | - | 0.31 (0.07-1.32) | - |
| Unadjusted | 3 - reopened | Women | 21 - 30 | Any case | 1 | 1.32 (1.06-1.66) | 1.67 (1.38-2.01) | 1.44 (1.19-1.74) | 1.30 (0.86-1.98) |
| Adjusted | 3 - reopened | Women | 21 - 30 | Any case | 1 | 1.33 (1.06-1.67) | 1.74 (1.44-2.11) | 1.50 (1.23-1.81) | 1.29 (0.85-1.97) |
| Unadjusted | 3 - reopened | Women | 21 - 30 | Hospitalisation | 1 | 0.80 (0.10-6.16) | 1.47 (0.44-4.93) | 0.34 (0.05-2.55) | 2.11 (0.25-18.11) |
| Adjusted | 3 - reopened | Women | 21 - 30 | Hospitalisation | 1 | 0.89 (0.11-7.08) | 1.80 (0.51-6.32) | 0.33 (0.04-2.68) | 1.76 (0.19-16.00) |
| Unadjusted | 3 - reopened | Women | 31 - 40 | Any case | 1 | 1.20 (0.98-1.47) | 1.47 (1.23-1.76) | 1.26 (1.05-1.51) | 1.32 (0.99-1.76) |
| Adjusted | 3 - reopened | Women | 31 - 40 | Any case | 1 | 1.32 (1.08-1.63) | 1.60 (1.33-1.93) | 1.33 (1.10-1.60) | 1.38 (1.03-1.85) |
| Unadjusted | 3 - reopened | Women | 31 - 40 | Hospitalisation | 1 | 0.62 (0.15-2.58) | 0.98 (0.30-3.21) | 1.93 (0.89-4.17) | 1.97 (0.57-6.87) |
| Adjusted | 3 - reopened | Women | 31 - 40 | Hospitalisation | 1 | 0.59 (0.14-2.54) | 0.93 (0.27-3.22) | 2.48 (1.11-5.57) | 2.05 (0.53-7.95) |
| Unadjusted | 3 - reopened | Women | 41 - 50 | Any case | 1 | 1.15 (0.91-1.46) | 1.58 (1.28-1.96) | 1.26 (1.03-1.54) | 1.22 (0.92-1.61) |
| Adjusted | 3 - reopened | Women | 41 - 50 | Any case | 1 | 1.09 (0.86-1.39) | 1.57 (1.25-1.96) | 1.24 (1.01-1.53) | 1.16 (0.87-1.56) |
| Unadjusted | 3 - reopened | Women | 41 - 50 | Hospitalisation | 1 | - | 0.55 (0.13-2.29) | 1.39 (0.54-3.58) | 2.60 (0.87-7.80) |
| Adjusted | 3 - reopened | Women | 41 - 50 | Hospitalisation | 1 | - | 0.41 (0.09-1.84) | 1.91 (0.71-5.14) | 2.09 (0.66-6.63) |
| Unadjusted | 3 - reopened | Women | 51 - 65 | Any case | 1 | 1.29 (1.00-1.66) | 1.77 (1.40-2.22) | 0.94 (0.73-1.22) | 0.92 (0.66-1.28) |

| Adjustment | Time period | Sex | Age group (years) | Outcome | Neither | Nursery/Primary or Nursery | Primary | Secondary | Teacher in other sector |
|---|---|---|---|---|---|---|---|---|---|
| Adjusted | 3 - reopened | Women | 51 - 65 | Any case | 1 | 1.20 (0.93-1.56) | 1.58 (1.24-2.01) | 0.91 (0.69-1.18) | 0.97 (0.69-1.36) |
| Unadjusted | 3 - reopened | Women | 51 - 65 | Hospitalisation | 1 | 0.58 (0.14-2.39) | 0.48 (0.12-2.01) | 0.89 (0.36-2.24) | 0.24 (0.03-1.73) |
| Adjusted | 3 - reopened | Women | 51 - 65 | Hospitalisation | 1 | 0.60 (0.14-2.57) | 0.48 (0.11-2.06) | 1.32 (0.51-3.39) | 0.39 (0.05-2.85) |
