## Supplementary material for "RISK OF HOSPITALISATION WITH COVID-19 AMONG TEACHERS COMPARED TO HEALTHCARE WORKERS AND OTHER WORKING-AGE ADULTS. A NATIONWIDE CASE-CONTROL STUDY": Statistical analysis plan

### Statistical analysis plan for study of the risk of COVID-19 in teachers in Scotland

Version 1.0

On the 20th of March 2020, nursery, primary and secondary schools closed as part of the Scottish response to the Covid-19 pandemic. All children, except vulnerable children and the children of keyworkers, stopped attending their usual school and continued their education via online resources with the support of parents and carers at home. Schools are expected to open to the majority of children on the 11th of August 2020.

To ensure that teachers, parents and carers, and pupils are confident in the safety of this process, and to inform policy-makers, it will be important to examine the impact of this change on COVID-19 infection. As part of the wider monitoring of COVID-19 in school settings taking place within PHS, we now propose to conduct a record-linkage study.

Record linkage involves combining different databases to generate new knowledge. A strength of record linkage studies is that data is collected in the same way for all groups being compared. This means that scientifically valid comparisons can be made; for example, rates of hospitalisation due to COVID-19 can be compared in teachers versus people who are not teachers.

The organisations which combined to form PHS have more than 30 years of expertise in performing record-linkage studies to monitor the health of the Scottish population. Recently, we used this expertise to link the Scotland-wide NHS human resources database to a range of health-related databases (eg databases on hospital admissions and testing for SARS-CoV2) in order to examine the risk of COVID-19 in NHS healthcare workers. A report on the findings of this analysis is due to be publicly released on the 5th of August and was made available to SG and the health boards via pre-release on the 29th of July.

We now propose to perform a similar analysis with teachers rather than healthcare workers (Figure 1). To that end, we require access to a database of teachers – and if available other education staff – in Scotland, which includes names, dates of birth and home postcodes. We propose to use these data in order to:-

1. Estimate the risk of hospitalisation, ICU admission and death due to COVID-19 among teachers in March and April 2020, and compare this with the general population.
2. Estimate the risk of SARS-CoV-2 infection, hospitalisation, ICU admission and death due to COVID-19 among teachers from August 2020 onwards, and compare this with the general population.

If the risks are found to be elevated in teachers, we will go on to estimate the risks in household members of teachers.

#### Figure 1 Overview of database linkages

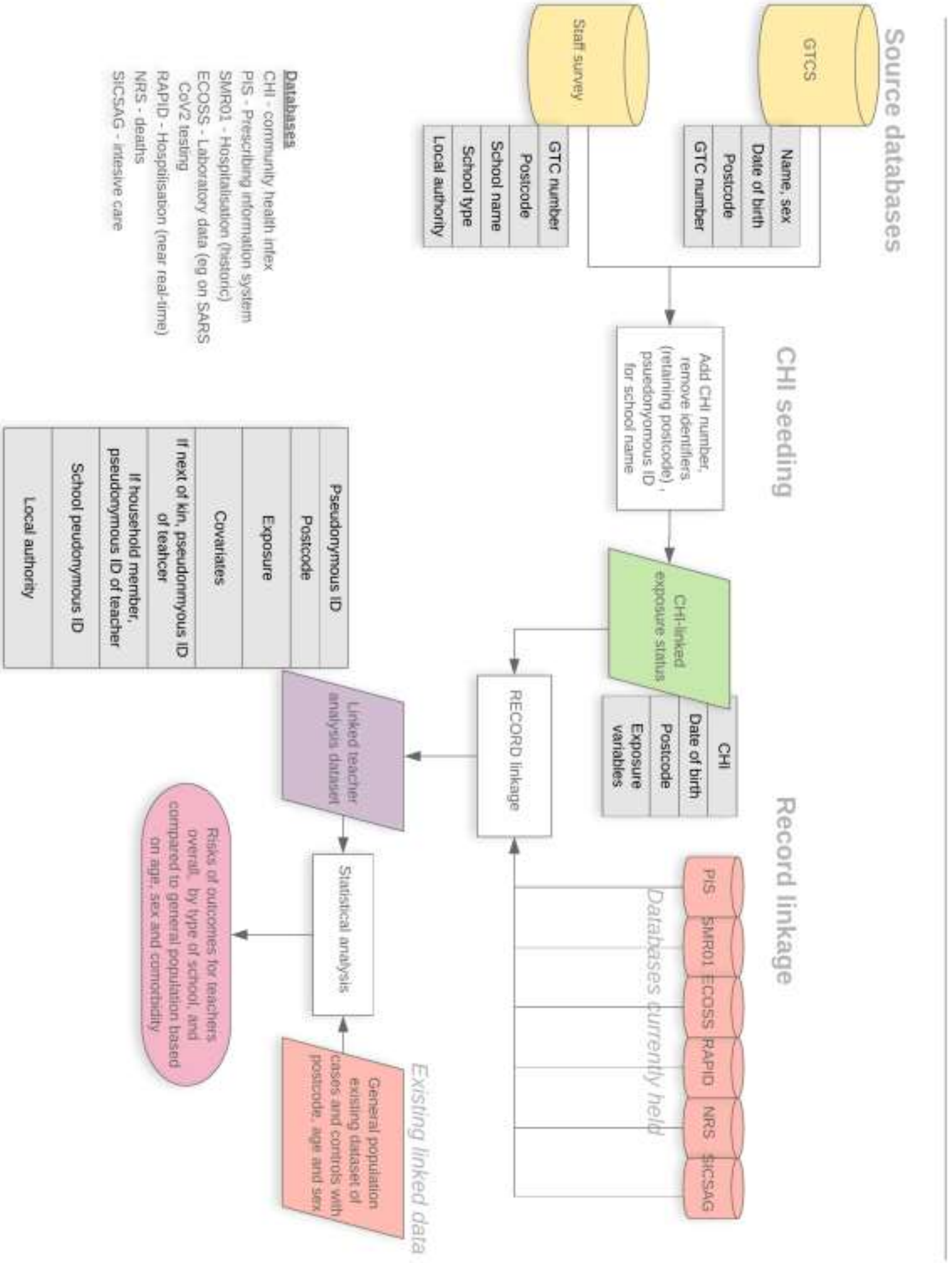

### Methods

#### Timeframe/cohort entry

Any teacher registered with the General Teaching Council on 1st March 2020 who is also identified on the staff survey as working in a primary or secondary school and as currently being in post. Teachers will be restricted to those aged between 18 and 65 (inclusive) as will general population comparators.

#### Setting

Teachers will be defined as secondary or primary school teachers based on the staff census. Where they are recorded as working in both settings they will be defined as secondary school teachers. Members of the working age population who are not teachers will be defined as general population comparators.

#### Covariates

In addition to age and sex, the covariates shown in Table 1 will be defined.

##### Table 1 Covariates

| Name | Definition |
|---|---|
| Chronic diseases | Pre-specified definitions based on SMR01 and pseudo-BNF codes. These are ischaemic heart disease, other heart disease, other circulatory disease, chronic kidney disease, chronic lower respiratory disease, neurological disorders, liver disease, immunodeficiency and immunosuppression, neoplasm, disorders of oesophagus, stomach and duodenum (see report on risk of COVDI-19 among healthcare workers for definitions). |
| Ethnicity | ONOMAP-derived ethnicity (https://www.onomap.org/). Teacher census reported ethnic background. |
| Scottish index of multiple deprivation | Area based measure of socio-economic deprivation |

#### Outcomes

We will use selected outcome definitions from the COVID19 case-control study already set-up within Public Health Scotland (REACT-COVID-19) (Table 2). Of these outcomes, “B Test positive and hospitalised” is the primary outcome. This was chosen to allow sufficient numbers while minimising ascertainment bias.

##### Table 2 Selected COVID-19 outcomes

| Outcome |
|---|
| A Test positive for SARS-COV2 |
| B Test positive and hospitalised |
| C Test positive and ICU |
| D Test positive, no ICU and died |
| E Test positive and ICU or died |
| F Test negative or no test, COVID19 on NRS death |

### Statistical analysis

#### Sample sizes and event numbers

In Scotland, 2152 people of working age were hospitalised within 28 days of their first positive test for SARS-CoV2, where that test was on or before the 1st of May 2020. There are 50,000 teachers in Scotland. Applying this rate (the denominator was 3,450,000) to the cohort of teachers (assuming the same age-sex distribution in teachers as in the general population) gives an expected count of 31.

Table 3 shows the statistical power, at alpha = 0.05, to detect a range of differences in rates between teachers and the general working age population based on there being 50,000 teachers currently working in schools in Scotland.

Table 3 Statistical power for detecting differences in rates

| N-fold higher rate in teachers | Statistical Power to detect difference |
|---|---|
| 1.2 | 0.19 |
| 1.3 | 0.34 |
| 1.4 | 0.53 |
| 1.5 | 0.70 |
| 1.6 | 0.84 |

#### Comparison of teachers with general population

We will compare rates in teachers to the general population via linkage to the existing case-control study – REACT-COVID-19. We will estimate rate ratios using conditional logistic regression models as these allow for the matching on age, sex and general practice area used in the case-control design. In further models we will additionally adjust for the covariates listed in Table 2. To accommodate clustering within schools, we will specify the school ID as a clustering variable within the survival::clogit function, which allows for correlation in the calculation of valid standard errors in the presence of clustering. This is a post-estimation correction – analogous to the approach used in generalised estimating equation models – which does not modify the effect estimate, but instead causes the standard errors to be inflated when there is evidence of correlation.

#### Additional analyses

In additional analyses, we will examine risks within teachers according to the setting (primary versus secondary school), and according to the level of contact time with pupils.

#### Presenting risk

Using outputs from the above models (and the baseline risks in the population), we will present estimates of the absolute risk of COVID-19 hospitalisation during the relevant period, alongside 95% confidence intervals. These risks will be presented according to age, sex, school, comorbidity and any other influential variables identified in the modelling.

#### Contingency analysis

The analysis is not possible without data from the General Teaching Council for Scotland (Figure 1). A restricted form of the analysis would be possible without the staff census data, however. This would involve estimating the risk for the total number of registered teachers of working age, regardless of whether these individuals are known to be in post. There are round 70,000 people registered as teachers with the General Teaching Council for Scotland, but only 50,000 current teachers. The remainder includes retired individuals (some of whom may be excluded based on age restrictions) and those who are no longer teaching but are still registered to teach (who we will not be able to exclude). Conversely the General Teaching Council database does not include all teachers working in the independent sector. As this work is urgent, it would be justified to undertake such an analysis, although it would systematically under-estimate any association between teaching and risk of COVID-19. Moreover, the additional analyses described above would not be possible without the staff census.
